# Hospital transmission of methicillin-resistant *Staphylococcus aureus* driven by addictive *mupA* plasmids

**DOI:** 10.64898/2026.07.24.26358837

**Authors:** Magdalena Podkowik, Ananyaa R. Welling, Somrita Dey, Alice Tillman, Gregory Putzel, Courtney Takats, Julian McWilliams, Stacey Bartlett, Nora Samhadaneh, Robert J. Ulrich, Kristine Rabii, Olufolakemi Olusanya, Caitlin Otto, Karl Drlica, Mila B. Ortigoza, Audrey Renson, Alejandro Pironti, Sarah Hochman, Bo Shopsin

**Affiliations:** Department of Medicine, Division of Infectious Diseases, NYU Grossman School of Medicine, New York, NY, USA; Department of Microbiology, NYU Grossman School of Medicine, New York, NY, USA; Antimicrobial-Resistant Pathogens Program, NYU Grossman School of Medicine, New York, NY, USA; Department of Population Health, NYU Grossman School of Medicine, New York, NY, USA; Clinical Microbiology and Diagnostic Immunology, NYU Grossman School of Medicine, New York, NY, USA; Public Health Research Institute, New Jersey Medical School, Rutgers University, Newark, NJ, USA; Department of Microbiology, Biochemistry & Molecular Genetics, New Jersey Medical School, Rutgers University, Newark, NJ, USA

**Author notes:** Correspondence: Bo Shopsin MD, PhD; 430 E 29^th^ St, New York, NY 10016, USA;, Sarah E. Hochman MD; 545 1^st^ Ave, SC1-173, New York, NY 10016, USA.

## Abstract

**Background:** Resistance to mupirocin, a cornerstone of *Staphylococcus aureus* decolonization, is a recognized cause of decolonization failure. Its role in hospital transmission is unknown.

**Methods:** We conducted genomic surveillance of >10,000 *S. aureus* isolates from adult patients at two urban hospitals where mupirocin decolonization is routine. Bacterial phenotypes and fitness were evaluated *in vitro* and in murine colonization models to interpret surveillance results.

**Results:** Genome sequencing identified 475 hospital transmission events; conventional surveillance detected none. The plasmid-mediated resistance determinant *mupA* (*ileS2*) was enriched eightfold in methicillin-resistant *S. aureus* (MRSA) relative to methicillin-susceptible strains. *mupA* was associated with nearly threefold greater hospital transmission, especially within healthcare-associated MRSA lineages. Surprisingly, multiple independently evolved inactivating mutations in the essential chromosomal gene *ileS1* co-occurred with *mupA*, creating plasmid addiction in which *mupA* became indispensable for bacterial survival. Plasmid carriage activated the stringent response and reduced colonization fitness, but also conferred collateral tolerance to disinfectants such as ethanol and peroxide. Although addiction further reduced *S. aureus* fitness, it increased plasmid transfer, promoting spread despite these costs. Unexpectedly, we found a mupirocin-dependent vulnerability to isoleucine limitation, revealing a potential strategy to target *mupA*-mediated resistance.

**Conclusions:** Hospital transmission of mupirocin-resistant MRSA is promoted by plasmids that create an evolutionary trap in which mupirocin use selects for bacterial dependence on costly resistance elements. These findings suggest that reducing mupirocin use alone is unlikely to eliminate resistance, underscore the need for genomic surveillance and resistance testing, and provide a framework for strategies to preserve mupirocin effectiveness.

**Major point:** This work shows that mupirocin resistance promotes hospital transmission of MRSA, identifies a previously unappreciated mechanism of plasmid addiction, and exposes a collateral vulnerability. These findings underscore the need for genomic surveillance and provide a framework to preserve mupirocin effectiveness.

## INTRODUCTION

Nasal mupirocin, often combined with chlorhexidine bathing, is widely used for decolonization and control of *Staphylococcus aureus*, especially methicillin-resistant strains (MRSA)[1–4]. These interventions help sustain recent reductions in healthcare-associated infections [5]. Plasmid-mediated mupirocin resistance is enriched in settings with extensive topical antimicrobial use [4]. As the principal decolonizing agent worldwide, the emergence and spread of resistance threaten infection prevention in vulnerable patients.

Mupirocin resistance arises through two mechanisms. Low-level resistance results from point mutations in the essential chromosomal isoleucyl-tRNA synthetase gene *ileS1*, whereas high-level resistance is mediated by *mupA*, which encodes an alternative isoleucyl-tRNA synthetase (*ileS2*) carried on plasmids [4, 6, 7]. We recently reported the emergence of a mupirocin-resistant community-acquired (CA-)MRSA lineage undergoing sustained community transmission in New York City [8, 9]. Whether hospital transmission is promoted by such plasmids and what evolutionary forces maintain them are unknown [10].

Here, we analyzed >10,000 *S. aureus* genomes collected between January 2022 and January 2025 to define the distribution, hospital transmission, and evolutionary consequences of *mupA*-carrying plasmids. Strong selection for *mupA* was evident in preferential hospital transmission of *mupA*-carrying strains and, more strikingly, in the repeated emergence (>100 independent times) of inactivating *ileS1* mutations that created plasmid addiction. In these strains, plasmid-encoded *ileS2* becomes essential for *S. aureus* survival. Broad biocide tolerance associated with plasmid carriage and increased conjugative transfer associated with addiction suggest that additional selective pressures reinforce plasmid maintenance and spread. These findings support proactive genomic surveillance to detect and mitigate the unintended evolutionary consequences of mupirocin use before they become entrenched. Importantly, analysis of resistance-fitness interactions revealed a collateral vulnerability: mupirocin synergized with exogenous isoleucine to re-sensitize *mupA*-positive strains, providing a rationale for adjuvant strategies to preserve mupirocin effectiveness and suppress MRSA in hospitals.

## METHODS

### Study isolates and procedures

*S. aureus* isolates were obtained from an institutional biobank at two tertiary-care hospitals within NYU Langone Health (Brooklyn and Manhattan; 444 and 813 beds). The biobank and decolonization procedures have been described previously [11, 12]. For the present study, we analyzed an expanded surveillance cohort comprising 10,321 sequenced isolates collected between January 2022 and January 2025, including 4,129 MRSA and 6,192 MSSA isolates. MRSA isolates were obtained from various clinical and admission screening cultures, whereas MSSA isolates were obtained from bloodstream infection and admission screening cultures only [12]. Adult patients admitted to medicine, oncology, transplant, and intensive care units underwent routine admission screening for nasal colonization. Patients colonized with MRSA or MSSA received twice-daily intranasal mupirocin and daily bathing with 2% chlorhexidine gluconate for 5 days. During the study period, admission screening compliance was 77.8%. Among colonized patients, 49% received at least one day of decolonization therapy and 10.8% received three or more days.

To identify putative progenitors of the *ileS1* mutant lineage E499, we screened a historical collection of 1,649 consecutive clinical MRSA isolates collected between 2017 and 2020 (pre-COVID-19 pandemic). These 604 isolates were obtained from bloodstream, respiratory, and wound infections in adult patients. The 31 *mupA*-positive CC8 isolates with closed genomes analyzed for chromosomal integration were also derived from this historical collection.

The study was approved by the NYU Langone Health Institutional Review Board (s24-01872). Additional details regarding strains, plasmids, primers, growth conditions, RNA sequencing, and phenotypic assays are provided in the Supplementary Methods.

### Genome sequencing and transmission detection

Whole-genome sequencing, assembly, quality control, bioinformatic analyses, and genomic surveillance–based transmission inference were performed as previously described for the institutional genomic surveillance program [11, 12]. Additional details, including transmission cluster definitions and linkage criteria, are provided in the Supplementary Methods.

## RESULTS

### Genomic Epidemiology of *mupA*-positive *S. aureus*

Susceptibility testing for mupirocin resistance is not routinely performed by most clinical laboratories because resistance is not included on automated testing platforms. We therefore used whole-genome sequencing to identify *mupA* across 10,321 *S. aureus* isolates collected through institutional genomic surveillance. Overall, *mupA* was detected in 11·8% of isolates, indicating a substantial reservoir of mupirocin resistance within the healthcare system. Mupirocin resistance was strongly enriched in MRSA. Although MSSA isolates were more numerous overall (6,192 versus 4,129), 85·8% (1,041) of *mupA*-positive isolates occurred in MRSA. Phylogenetic analysis revealed that *mupA* within MRSA was concentrated in CC8 and, to a lesser extent, CC5. These lineages represent the predominant community- and healthcare-associated MRSA lineages, respectively, in New York City and the United States [13–15]. Together, these lineages accounted for 96·7% of *mupA*-positive MRSA, whereas the remainder of *mupA*-positive isolates occurred in sporadic isolates and small clusters. We therefore focused subsequent analyses on CC8 and CC5.

*mupA* was distributed across multiple independent branches of both the CC5 and CC8 phylogenies (Fig. 1A), consistent with repeated horizontal acquisition, not single ancestral expansion. Nevertheless, several large clonal expansions were evident, particularly within CC8.

**Figure 1.**
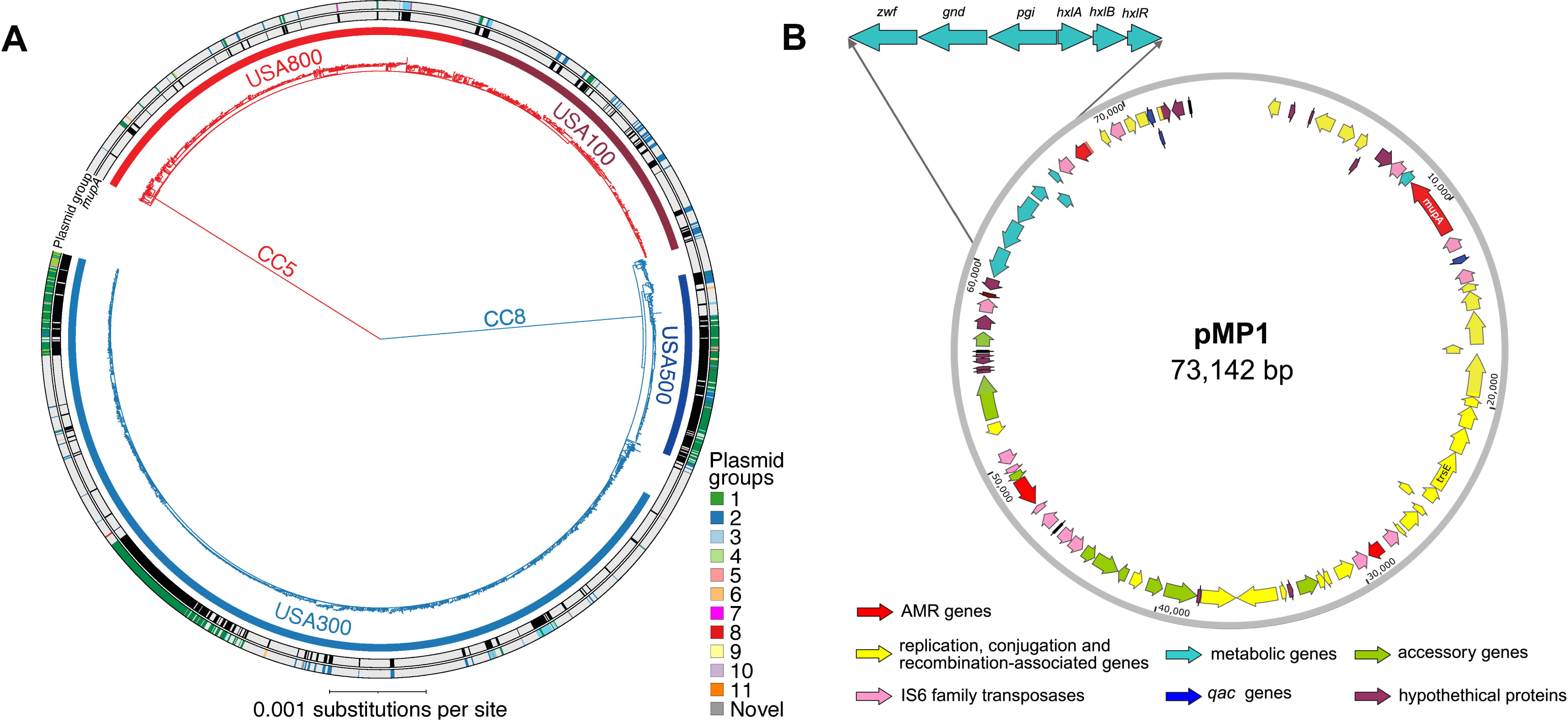
Distribution and characteristics of *mupA*-containing plasmids among epidemic MRSA lineages. **(A)** Maximum-likelihood phylogeny of CC5 and CC8 MRSA isolates from the surveillance cohort (*n* = 1,945), midpoint rooted. Clonal complexes are indicated on the tree. For each patient, a single representative susceptible or resistant isolate was included (<10 patients contributed both a *mupA*-positive and *mupA*-negative isolate). Outer ring, presence of *mupA* (red) or absence of *mupA* (gray). Middle ring, MOB-suite–imputed plasmid groups. Inner ring, major epidemic lineages (CC5, USA100 and USA800; CC8, USA300 and USA500), assigned by *in silico* detection of lineage-defining genetic elements and color-coded as indicated. Lineages were defined by the presence of characteristic genetic markers in >70% of isolates within each clonal complex, as described.[12] **(B)** Representative *mupA*-containing conjugative plasmid pMP1 (73,142 bp), illustrating Group 1, one of the three major *mupA*-associated plasmid groups identified in the surveillance cohort. pMP1 carries mupirocin resistance determinant *mupA* together with multiple insertion sequences and transposons, additional antimicrobial resistance determinants including the antiseptic resistance gene *qacC*, a phosphoenolpyruvate phosphatase (PPP) shunt metabolic operon, and genes involved in plasmid conjugation and mobilization. Representative plasmids from the three major plasmid groups and their structural relationships are shown in Supplementary Fig. 1.

Assembly of a subset of complete plasmids revealed several structurally distinct plasmid groups (representative plasmid in Fig. 1B; group comparisons in Supplementary Fig. 1). These complete plasmids informed plasmid classification across the surveillance cohort (Fig. 1). Three major plasmid groups (Groups 1–3) accounted for >91% of *mupA*-positive isolates, with Group 1 representing 60%. Thus, *mupA* dissemination was dominated by a small number of plasmid groups. Mobility predictions of the reference plasmids corresponded closely with the major structural groups and were consistent with a recent regional survey (Supplementary Fig. 1) [16]; representative Group 1 plasmids were conjugative, Group 2 included mobilizable and non-mobilizable plasmids, and Group 3 plasmids were mobilizable. Conjugative plasmids contained a *tra* region homologous to those of the *S. aureus* resistance plasmids pUSA03 and pSK41 [7, 17].

Plasmids exhibited extensive mosaicism, consistent with recombination and exchange of accessory genetic elements (Supplementary Fig. 1). In addition to *mupA*, these plasmids encoded multiple accessory features, including antimicrobial resistance determinants, transposases, and insertion sequences. Notably, 53·7% of *mupA*-positive strains also carried chlorhexidine resistance genes, establishing dual resistance to both hospital decolonization agents, consistent with previous reports [8, 18, 19]. Some plasmids also carried a pentose phosphate pathway locus arranged in a non-native, operon-like configuration (Fig. 1B). Mapping plasmid structural groups onto the chromosomal phylogeny revealed both multiple plasmid groups within individual lineages and identical plasmid groups across lineages (Fig. 1A). These findings confirm that *mupA* plasmids spread through a combination of vertical clonal expansion and repeated inter-lineage transfer.

### *mupA* is Associated with Increased Patient-to-patient Transmission

Patients harboring *mupA*-positive isolates differed in several previously identified risk factors for *S. aureus* colonization and transmission, including greater hospital exposure (Supplementary Table 1) [12]. To determine whether *mupA* was associated with transmission, we used a previously described genomic-epidemiologic framework (Supplemental Methods) [11, 12]. Compared with *mupA*-negative strains, *mupA*-positive strains had a 10·5 percentage point higher transmission rate (95% CI 8·4 – 12·7); this association was robust to propensity score adjustment for lineage, hospitalization history, antibiotic exposure, and demographic factors (adjusted difference 11·3 percentage points, 95% CI 5·8–16·9; Supplementary Table 2). Consistent with transmission contributing to infection risk [20], *mupA* was also enriched among hospital-onset infections compared with admission colonizing isolates (17·0% vs 10·2%).

The transmission advantage associated with *mupA* was observed across both MRSA and MSSA but was most pronounced in MRSA lineages (Fig. 2, Supplementary Table 3). MSSA increased from 2·1% to 7·0% with *mupA* but remained low overall; mupirocin-resistant MSSA transmitted at rates comparable to mupirocin-susceptible MRSA. In CC5 MRSA, transmission increased from 12·9% to 26%, and in CC8 MRSA the increase was from 7·4% to 14·6%. Thus, although *mupA* was most prevalent in CC8, its association with transmission was greatest in CC5, the predominant healthcare-associated MRSA lineage.

**Figure 2.**
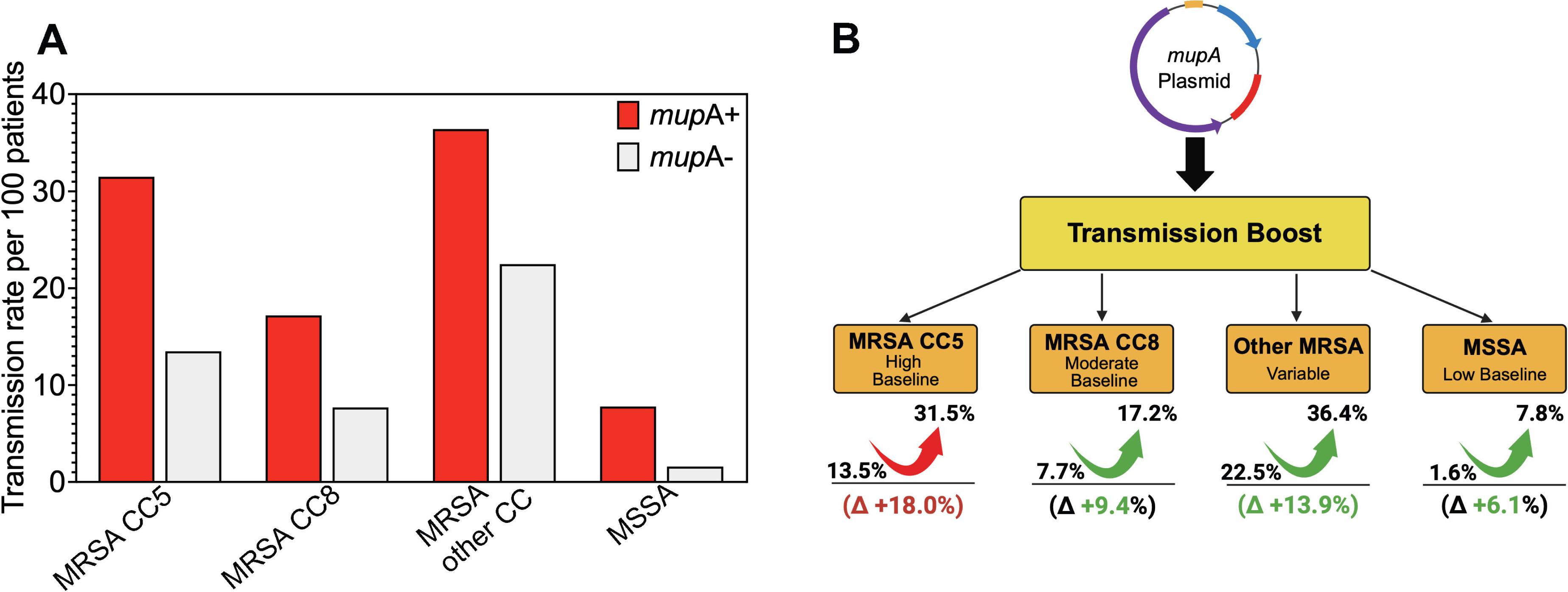
*mupA* carriage is associated with increased transmission across diverse *S. aureus* lineages. **(A)** Transmission rates among *mupA*-positive (red) and *mupA*-negative (gray) isolates stratified by MSSA, MRSA CC5, MRSA CC8, and other MRSA clonal complexes. Bars indicate the proportion of isolates within each category linked to at least one transmission event. Transmission events were defined by combined genomic and epidemiologic linkage, including direct and indirect epidemiologic connections between patients carrying isolates separated by ≤20 single-nucleotide variants (Methods). In multivariable analyses, *mupA* carriage remained independently associated with transmission after adjustment for host and epidemiologic factors (Supplementary Table 2). **(B)** Schematic summary of the absolute increase in transmission associated with *mupA* carriage across lineage groups with differing baseline transmission frequencies. Values shown are derived from panel A and are presented to conceptually illustrate the relationship between baseline transmission and the magnitude of the *mupA*-associated transmission advantage. Numerical values, confidence intervals, and statistical comparisons are provided in Supplementary Table 3.

Chromosomal *ileS1* point mutations conferring low-level mupirocin resistance (V588F, V631F, G593V)[21] were not associated with increased transmission. The consistency of the *mupA* effect across genetic backgrounds and patient populations, its persistence after statistical adjustment, and the absence of effect from chromosomal resistance mutations support a role for plasmid-mediated mupirocin resistance in promoting hospital transmission.

### Recurrent Evolution of Plasmid Addiction Through Chromosomal *ileS1* Inactivation

Unexpectedly, analysis of *ileS1* revealed widespread inactivation of this essential chromosomal isoleucyl-tRNA synthetase among *mupA*-positive isolates; no loss-of-function mutations were identified in mupA-negative isolates. We identified 105 distinct loss-of-function mutations, consisting of frameshifts and truncations (Fig. 3A). Most mutations were observed in only one or a few isolates; however, several underwent substantial clonal expansion, including one lineage involving >155 patients. Isolates carrying the same inactivating *ileS1* mutation clustered within monophyletic groups, indicating that recurrently observed mutations reflected clonal expansion rather than parallel evolution (Fig. 3B; Supplementary Fig. 2).

**Figure 3.**
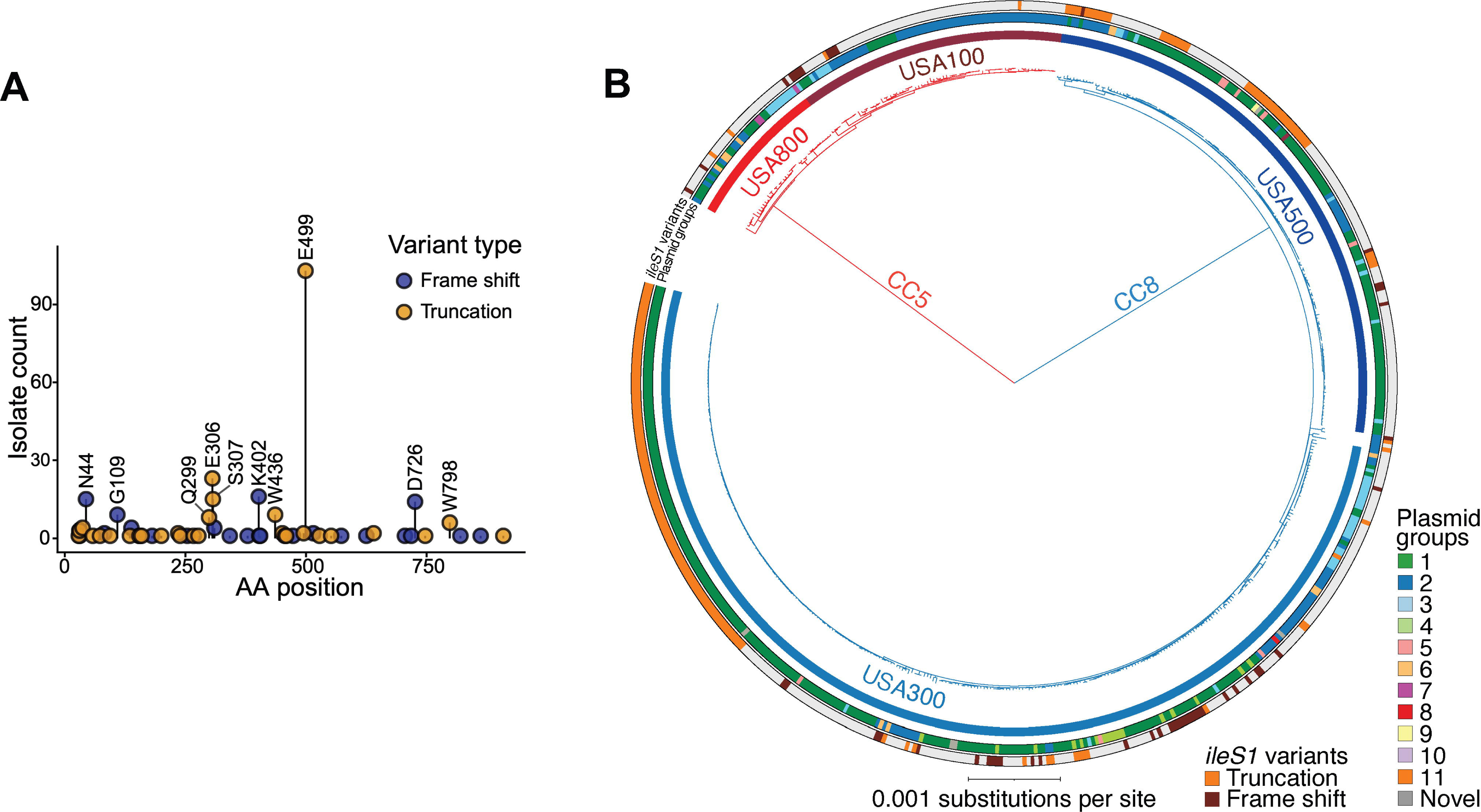
Diversity and phylogenetic distribution of chromosomal *ileS1* inactivation mutations among *mupA*-containing epidemic MRSA lineages. (A) Summary of insertion and deletion mutations predicted to inactivate chromosomal *ileS1* among *mupA*-positive MRSA isolates (*n* = 103). Each point represents a unique mutation identified among patient isolates, plotted according to its position within *ileS1*. Mutations are classified as frameshift (blue) or truncating (gold) variants. Amino acid positions are labeled using single-letter amino acid abbreviations. One representative isolate per patient was included in the analysis. The E499 truncation was the most frequently observed mutation. **(B)** Maximum-likelihood phylogeny of *mupA*-positive CC5 and CC8 MRSA isolates from the surveillance cohort (*n* = 574), midpoint rooted. Isolates correspond to the *mupA*-positive subset shown in Fig. 1A and are limited to a single representative isolate per patient. Clonal complexes, lineage assignments, and plasmid groups are annotated as described in Fig. 1. The outer ring indicates chromosomal *ileS1* inactivation mutations (frameshift or truncation). The largest cluster of *ileS1* mutants carries the E499 truncation and is further characterized in Supplementary Fig. 2.

Addiction (*leS1* inactivation–mediated dependence on *mupA*) was most common in CC8 and in plasmid groups associated with this lineage, reflecting the high prevalence of *mupA* and expansion of addicted clones. Thus, plasmid addiction both reflects MRSA population structure and contributes to its evolution.

Because chromosomal integration events are largely invisible in short-read surveillance data, we screened all available closed CC8 genomes from our collection. Among 31 *mupA*-positive isolates, chromosomal integration was identified in 6 (19·4%). All six occurred among the seven addicted isolates (85·7%), involving three independent addiction variants, whereas none of the 24 non-addicted isolates showed chromosomal integration. Thus, chromosomal acquisition of *mupA* may be an additional outcome of plasmid addiction.

### Fitness Costs of *mupA* Plasmids Drive Selection for Addiction

To determine whether *ileS1* inactivation conferred a fitness advantage, we introduced an *ileS1* addiction mutation identified in a clinical isolate into a laboratory strain carrying a *mupA* plasmid. Addiction increased neither resistance nor growth in the presence of subinhibitory mupirocin concentration (Supplementary Fig. 3). Instead, addicted strains grew more poorly than their plasmid-carrying parental counterparts, indicating that addiction imposes a burden rather than enhancing resistance.

These findings suggest that addiction is selected not to improve bacterial growth but to stabilize plasmids that would otherwise be lost. Consistent with this interpretation, plasmid carriage produced no measurable defect *in vitro* across a range of growth conditions, but it imposed substantial costs *in vivo*. In a murine nasal colonization and transmission model (Fig. 4A), introduction of the plasmid into a CC8 background, where addiction is common, significantly reduced both colonization and transmission (Fig 4B, Supplementary Fig. 4A). By contrast, in CC5, where addiction is less common, plasmid carriage had minimal or slightly beneficial effects (Fig. 4C, Supplementary Fig. 4B). Introduction of an addicted chromosomal background further reduced transmission (Supplementary Fig. 5), demonstrating that addiction can persist despite exacerbating plasmid-associated fitness costs. Together, these findings support the idea that fitness costs promote selection for plasmid-stabilizing addiction and may contribute to its differential prevalence among MRSA lineages.

**Figure 4.**
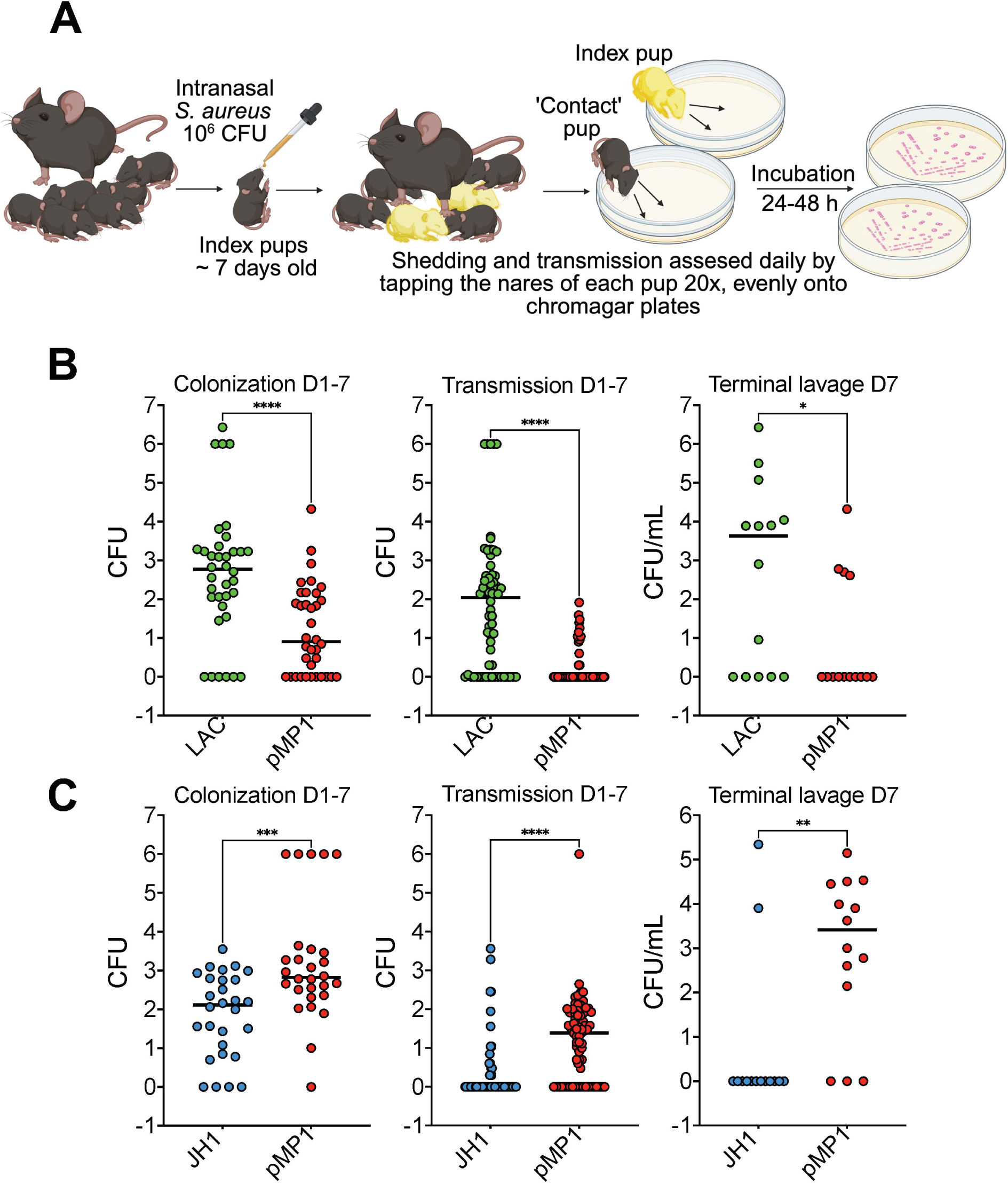
Effect of the *mupA* plasmid on colonization and transmission in a neonatal murine model. **(A)** Experimental schematic. Neonatal mice (4–7 days of age) were intranasally inoculated with 10^6^ CFU of the indicated *S. aureus* strain. Inoculated index pups were returned to their litters and cohoused with uninoculated littermates to assess transmission. Colonization was monitored daily by gently tapping the nares of each pup onto selective agar plates. At experimental endpoints, pups and dams were euthanized and upper respiratory tract colonization was quantified by retrograde nasal lavage and CFU enumeration. **(B)** Colonization and transmission of the CC8 strain LAC ± pMP1 (BS1158 and BS1963, respectively). **(C)** Colonization and transmission of the CC5 strain JH1 ± pMP1 (BS1231 and BS2045, respectively). Left, cumulative nasal colonization burden among inoculated index pups measured over days 1–7 post-inoculation. Middle, cumulative transmission burden among uninoculated contact pups measured over the same interval. Right, terminal upper respiratory tract colonization quantified by retrograde nasal lavage at the experimental endpoint. In **panel B**, green indicates the parental strain and red the *mupA*-plasmid–containing strain. In **panel C**, blue indicates the parental strain and red the *mupA*-plasmid–containing strain. Each symbol represents an individual animal. For longitudinal colonization measurements (left and middle panels), each symbol represents the cumulative CFU recovered from a single animal over the 7-day observation period. Whenever no *S.aureus* was recovered a value of 1 CFU was assigned. Horizontal bars indicate medians. All Y axes are in log scale. Daily colonization and transmission kinetics for the experiments shown in panels B and C are presented in Supplementary Fig. 4.

### *mupA* Plasmids/Addiction Impose Dependence on the Stringent Response

Because aminoacyl-tRNA synthetases operate near saturation, even small activity reductions impose steep fitness costs [22]. Reduced charging efficiency of the *mupA*-encoded isoleucyl-tRNA synthetase may therefore elevate uncharged tRNA levels and activate the stringent response via production of (p)ppGpp, a nutrient-stress pathway triggered by accumulation of uncharged tRNA that suppresses growth and biosynthesis [23]. Experimental disruption of the stringent-response is complicated in *S. aureus* because the RelA/SpoT homolog (RSH), which mediates (p)ppGpp synthesis and hydrolysis, is essential for viability [24]. Therefore, we evaluated growth in the presence of relacin, an inhibitor of (p)ppGpp synthesis, to test whether *mupA*-carrying strains exhibit increased dependence on stringent-response signaling, as predicted for aminoacyl-tRNA synthetase limitation [22]. Relacin impaired growth of plasmid-carrying and addicted strains, but it had little effect on plasmid-free, parental strains (Fig. 5). Although relacin exhibits low affinity for Rel and may have off-target effects [25], these findings support the idea that *mupA* plasmids increase dependence on the stringent response. This dependence provides a mechanistic explanation for both the fitness cost of plasmid carriage and selection of addicted *ileS1* mutants.

**Figure 5.**
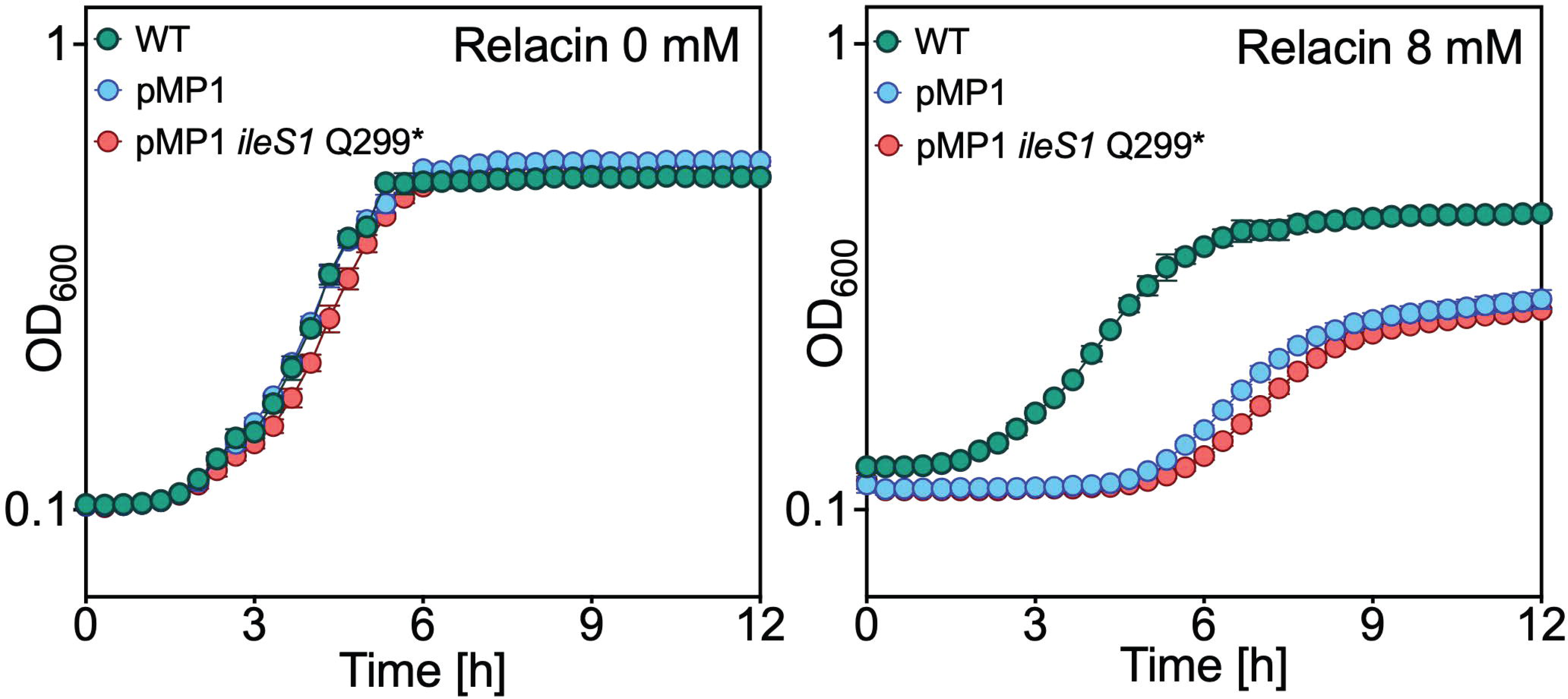
Effect of Relacin on growth of *mupA*-containing strains with and without chromosomal *ileS1* inactivation. **(A)** Growth of *S. aureus* LAC wild-type (WT; BS1158), the isogenic *mupA* plasmid-containing derivative (pMP1; BS1963), and the addicted pMP1 *ileS1* Q299* mutant (BS2039) in TSB. Optical density at 600 nm wavelength was recorded using a BioTek LogPhase 600 microbiology reader. **(B)** Growth of the same strains in TSB as in A, supplemented with Relacin (at 8 mM). Data represent means ± SD from biological replicates (*n* = 3).

Because stringent-response activation promotes biofilm formation [26], we examined this phenotype. Addicted strains, but not their plasmid-carrying parental counterparts, exhibited increased biofilm formation compared with plasmid-free controls (Fig. 6A). Because conjugative transfer is enhanced within biofilms [27], we next tested whether addiction influences plasmid transfer. Conjugation frequencies were higher in addicted strains than in their plasmid-carrying parental strains (8·88e-7 vs 2·31e-7 (mean values); Fig. 6B). Thus, addiction not only ensures vertical inheritance but it also promotes horizontal dissemination, thereby expanding the reservoir of transmissible resistance [28].

**Figure 6.**
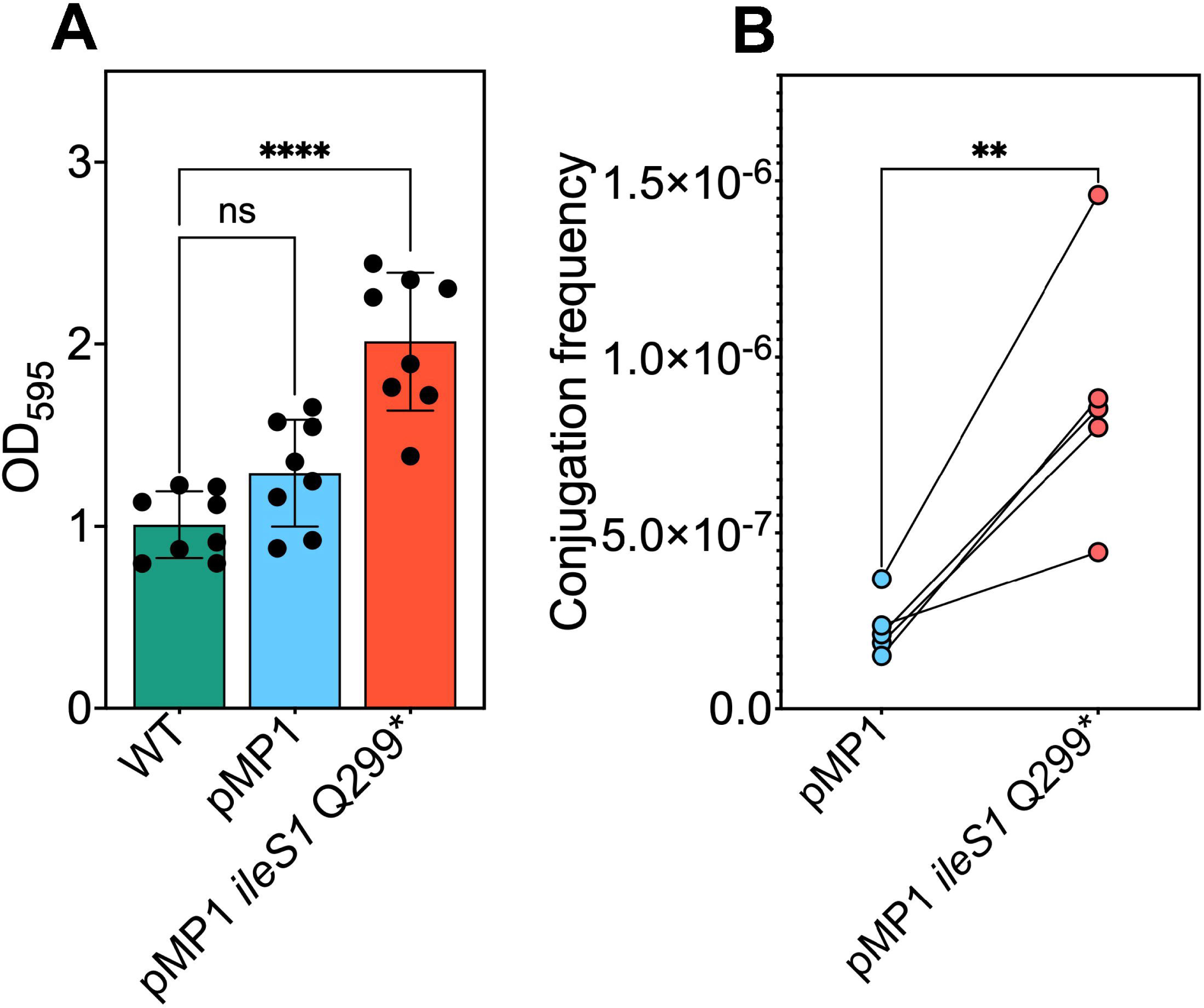
Effect of the *mupA* plasmid and chromosomal *ileS1* inactivation on biofilm formation and plasmid transfer. **(A)** Biofilm production *S. aureus* LAC wild-type (WT; BS1158), the isogenic *mupA* plasmid-containing derivative (pMP1; BS1963), and the addicted pMP1 *ileS1* Q299* mutant (BS2039). Biofilm biomass was quantified by crystal violet staining following 24 h of static growth and measured at OD_600_. Bars represent mean ± SD. Data are from 4 independent experiments. Statistical significance was determined by one-way ANOVA with Dunnett’s multiple-comparisons test; ****P ≤ 0.0001; ns, not significant. (**B**) Conjugative transfer of pMP1 by filter mating. Transfer frequencies were determined using either the parental LAC recipient strain (blue) or an isogenic pMP1-containing *ileS1* Q299* recipient strain (red). Each point represents an independent experiment (*n* = 5); lines connect paired replicate experiments. Conjugation frequencies are expressed as transconjugants per recipient. Statistical significance was determined by unpaired Student’s t test; **P ≤ 0.01.

### Stringent-Response Dependence Creates a Therapeutic Vulnerability

Expression analysis of selected stringent-response genes in biofilm populations was consistent with activation of stringent-response pathways in both *mupA* plasmid-carrying and addicted strains (Supplementary Fig. 6), as evidenced by increased expression of oligopeptide transport genes (*oppA*, *oppC*), branched-chain amino acid (BCAA) biosynthesis genes (*ilvB*, *leuA*, *leuB*), purine salvage (*xpt*), and reduced expression of ribosomal genes (*rplR*, *rplV*), purine biosynthesis (*purH)*, and metabolism (*sucA*, *atpA, gltB*) [29, 30]. The addicted strain generally showed larger effect sizes across several markers, potentially explaining its association with increased biofilm formation. No change in *rsh* expression was observed, consistent with prior work showing that stringent-response transcriptional programs can persist after adaptation to sublethal stress without increased *rsh* transcription [31, 32]. Reduced translational capacity and impaired energy metabolism provide a potential mechanistic basis for the fitness cost of plasmid carriage.

The stringent-response dependence imposed by *mupA*, and more generally by mupirocin [33], suggested a vulnerability arising from the interaction between stringent-response signaling and CodY-mediated regulation. In *S. aureus*, intracellular pools of BCAA are integrated with stringent-response signaling through the global regulator CodY [34–37]. When BCAA are abundant, CodY represses genes involved in amino acid biosynthesis and stress adaptation; depletion of these metabolites relieves repression and activates these pathways. Thus, exogenous isoleucine can uncouple this regulatory relationship by reactivating CodY even when stringent-response signaling persists [36–38]. Among BCAA, isoleucine is the most potent mediator of this effect, restoring CodY-dependent repression despite continued metabolic stress [36, 37]. This creates a regulatory mismatch in which stringent-response signaling remains active while CodY suppresses the biosynthetic programs needed for growth.

This interaction has limited consequences under nutrient-replete laboratory conditions, where amino acids are abundant. However, the *in vivo* environment differs substantially. Nasal secretions, the primary ecological niche for *S. aureus* colonization, are markedly deficient in free amino acids, especially isoleucine [39]. Under these conditions, mupirocin stringent-response activation occurs in the setting of amino-acid limitation, where relief of CodY repression facilitates biosynthesis and growth. Exogenous isoleucine would be expected to disrupt this adaptive response by restoring CodY despite ongoing stringent-response signaling, with consequences for growth [36, 37].

Consistent with this model, moderate concentrations of isoleucine alone modestly impaired growth with both plasmid-free and plasmid-carrying strains grown in defined medium lacking BCAA (Fig. 7A). However, when combined with subinhibitory mupirocin, which further impairs isoleucyl-tRNA charging and amplifies stringent-response signaling, even lower doses of exogenous isoleucine produced near-complete growth collapse (Fig. 7B). Under these conditions, either agent alone had no impact. This synergistic interaction demonstrates a vulnerability in *mupA*-containing strains during mupirocin exposure, allowing resistance to be sensitized without directly inhibiting the resistance determinant.

**Figure 7.**
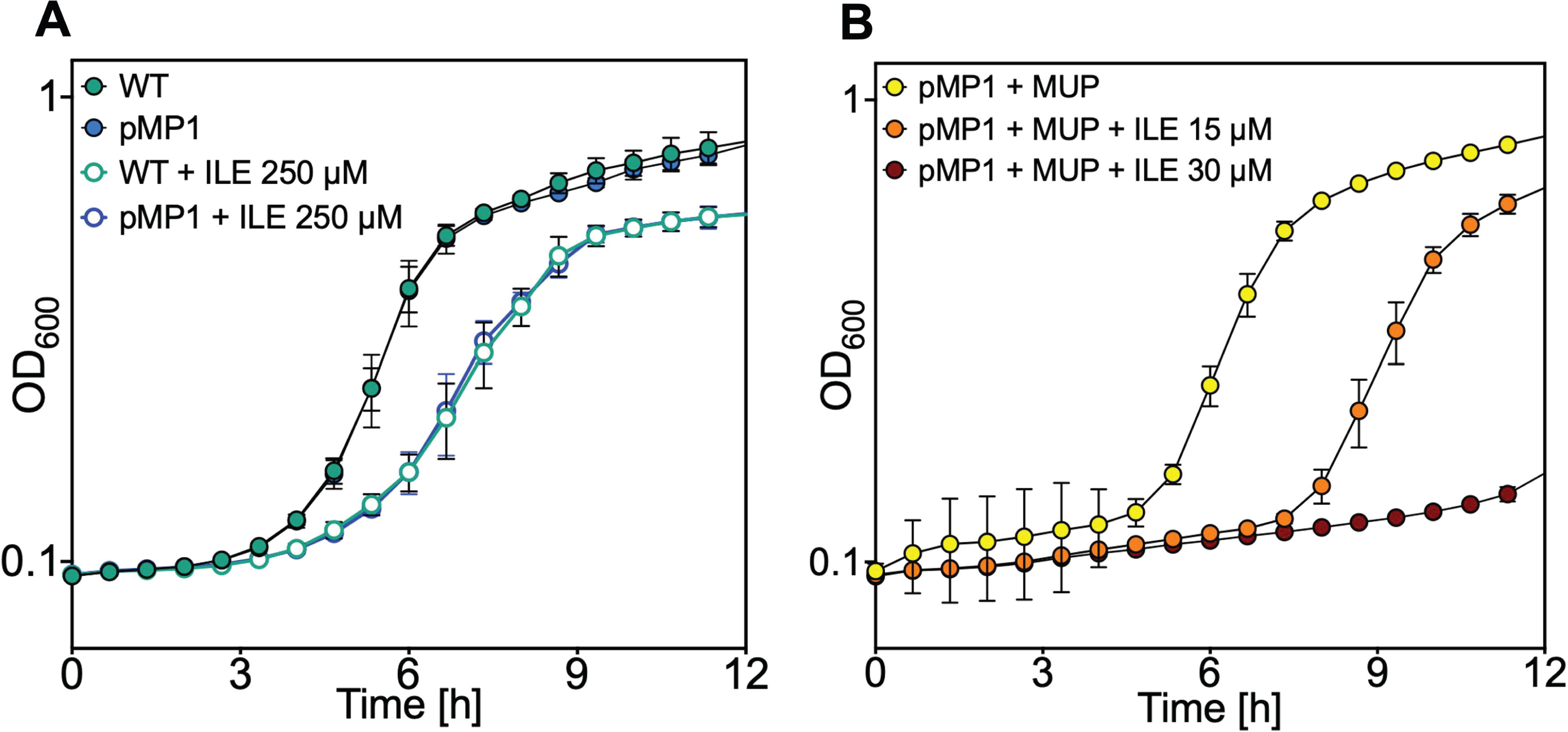
Exogenous isoleucine synergizes with subinhibitory mupirocin to impair growth of *mupA*-containing strains. **(A)** Growth of *S. aureus* LAC wild-type (WT; BS1158) and the isogenic *mupA* plasmid-containing derivative (pMP1; BS1963) in TSB medium, with or without supplementation with isoleucine at the indicated concentration. **(B)** Growth of the pMP1 strain in BCAA-free RPMI medium in which growth depends on endogenous BCAA biosynthesis, supplemented with subinhibitory mupirocin (100 μg/mL) alone or in combination with isoleucine at the indicated concentration. Growth was recorded using a BioTek LogPhase 600 microbiology reader, measuring OD_600_ at 40-min intervals. Data represent means ± SD from biological replicates (*n* = 3).

Identifying vulnerabilities is important because inhibition of tRNA synthetases is known to induce broad tolerance to disinfectants [40]. The resulting stringent response suppresses the reactive oxygen species surge induced by bactericidal agents, thereby reducing killing and promoting survival under biocidal stress. We therefore tested whether *mupA* plasmids altered tolerance to common biocidal agents after growth in BCAA-deficient medium, conditions that approximate the amino-acid limitation of nasal secretions. Plasmid-carrying strains exhibited a 1–2 log reduction in killing following exposure to multiple disinfectants, including ethanol, 2-propanol, and hydrogen peroxide, versus a plasmid-free parental strain (Fig. 8). In contrast, addicted strains did not show further increases in tolerance beyond that conferred by the plasmid alone. Thus, *mupA* plasmids confer broad biocide tolerance.

**Figure 8.**
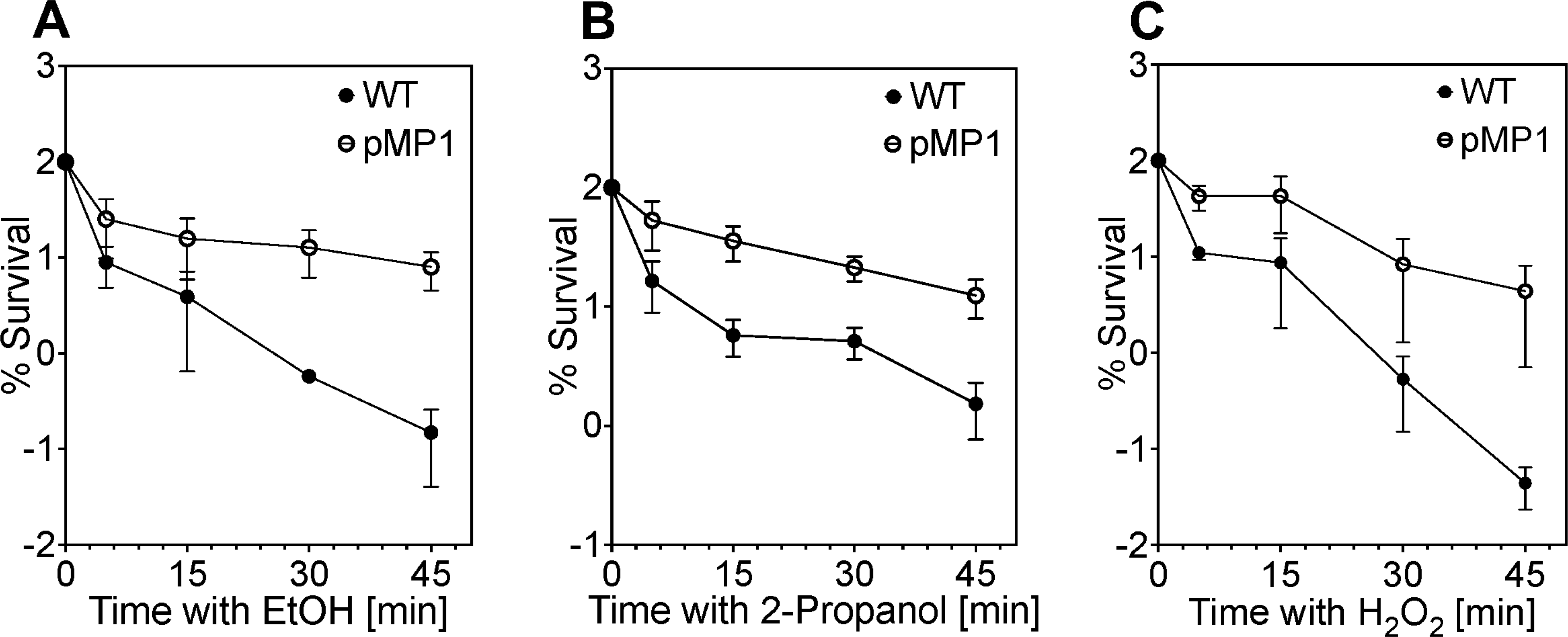
*mupA* plasmid increases broad-spectrum survival during disinfectant-mediated bacterial killing. Kinetics of killing by ethanol, 2-propanol and H_2_O_2_. Overnight cultures of *S. aureus* LAC (WT; BS1158) and the isogenic *mupA* plasmid-containing derivative (pMP1; BS1963) grown in BCAA-free RPMI medium, in which growth depends on endogenous BCAA biosynthesis, were diluted (OD_600_ ≈ 0.05) into fresh tryptic soy broth (TSB) and grown with orbital shaking (180 rpm) to early exponential phase (OD_600_ ≈ 0.15). Cultures were then exposed to **(A)** ethyl alcohol (20%), **(B)** 2-propanol (13%), or **(C)** hydrogen peroxide (H_₂_O_₂_; 20 mM) for the indicated times. Survival was quantified by CFU enumeration and expressed as log_₁₀_ percent survival relative to CFU at time of disinfectant addition. Data represent means ± SD from biological replicates (*n* = 3). No differences in MICs for ethanol, 2-propanol, or H_2_O_2_ were detected between the parental and *mupA* plasmid-containing strains.

## DISCUSSION

The present study of hospital transmission associated with mupirocin resistance uncovered a previously unappreciated mechanism by which *S. aureus* becomes locked into plasmid carriage, stabilizing mupirocin resistance. In these cases, mutational inactivation of the essential chromosomal *ileS1* gene renders bacterial survival dependent on plasmid-encoded *ileS2*, creating plasmid-host interdependence that ensures retention even in the absence of mupirocin selection. This mechanism follows the logic of toxin-antitoxin systems—plasmid loss is more costly than retention—but dependence arises through essential-gene replacement rather than post-segregational killing [41], precluding survival of plasmid-free bacteria. Addiction emerged repeatedly, underscoring strong selection to stabilize resistance plasmids. Entrenchment of addicted lineages within MRSA populations would pose a public-health threat.

Chromosomal integration of *mupA* among addicted strains is consistent with the expectation that beneficial accessory genes may ultimately be captured by the chromosome [42]. However, unlike classical models, addiction creates dependence on a plasmid-borne function through loss of the chromosomal homolog. Thus, addiction may represent another route by which plasmid-borne genes become assimilated into the host genome.

Addiction increased biofilm formation and conjugative transfer of *mupA*. Thus, addiction not only ensures vertical inheritance but also promotes horizontal dissemination, generating a plasmid reservoir that spreads resistance within and across MRSA lineages. Consistent with these experimental findings, phylogenetic analyses revealed expansion of addicted clones and repeated acquisition of *mupA*-containing plasmids (Fig. 2A, Supplementary Fig. 2). Together, these observations support a role for addiction in dissemination of *mupA*.

Whereas addiction promotes persistence of *mupA* within MRSA populations, *mupA* itself was associated with an increased likelihood of hospital transmission. This transmission advantage was strongest in MRSA and greatest within CC5, the predominant healthcare-associated MRSA lineage, and smallest in MSSA. Thus, *mupA* preferentially enhanced transmission among strains that already exhibit the highest baseline transmission potential in healthcare settings (Fig. 2, ref.[12]). Rather than creating successful lineages, *mupA* appears to amplify the fitness of strains already adapted to transmission. The molecular basis of this effect is unknown.

The greater absolute transmission advantage associated with *mupA* in healthcare-associated CC5 MRSA, despite the higher prevalence of *mupA* in CC8, suggests that healthcare transmission is only one component of the selective environment shaping *mupA* dissemination. Indeed, prior work identified the emergence of a mupirocin-resistant CC8 lineage undergoing community transmission [8] and showed that hospital transmission risk reflects interactions between host epidemiology and bacterial lineage [12]. Consistent with this interaction, the present study found that CC8, but not CC5, incurred an *in vivo* fitness cost associated with *mupA* carriage and exhibited a higher burden of addiction, suggesting stronger selection for plasmid stabilization. The relative contributions of clonal spread, horizontal plasmid transfer, and plasmid addiction to sustaining *mupA* carriage when selective pressure is reduced likely differ between CC5 and CC8, thereby promoting persistence in their primary epidemiologic settings [43]. Thus, strategies based solely on restricting mupirocin use may be insufficient in both settings.

*mupA* plasmids conferred increased survival under biocide pressure, including ethanol, isopropanol, and chlorhexidine. Increased tolerance to these agents could enhance environmental persistence and opportunities for transmission, potentially contributing to the increased transmission of *mupA*-carrying strains observed in our surveillance data. Together with additional metabolic and resistance determinants encoded by *mupA* plasmids, these findings suggest that the success of *mupA* plasmids may reflect broader adaptive benefits beyond resistance to mupirocin alone.

Exogenous isoleucine synergized with mupirocin to inhibit growth of *mupA*-containing strains under BCAA-limited, nasal-like conditions. A likely explanation, modeled in Supplementary Fig. 7, is that by impairing isoleucyl-tRNA charging, mupirocin reduces utilization of intracellular isoleucine, thereby increasing its effective availability. As a result, even low concentrations of exogenous isoleucine may be sufficient to maintain CodY-mediated repression of amino acid biosynthetic pathways. The resulting regulatory mismatch suppresses biosynthetic programs required for BCAA synthesis, creating a conditional vulnerability that culminates in growth collapse.

Several limitations warrant consideration. Further genomic and experimental studies are needed to confirm the predicted low charging efficiency of *mupA*-encoded IleS2, demonstrate that stringent-response activation underlies plasmid-associated fitness costs, determine why these costs are particularly pronounced in CC8, and identify compensatory mutations that mitigate conflicts between plasmid carriage and the host chromosome in successful addicted lineages [44]. Additional *in vivo* studies, including colonization and transmission models, are underway to determine whether the isoleucine-mediated sensitization observed *in vitro* can be translated into an effective decolonization strategy. Finally, our epidemiologic analyses were limited to two hospitals within a single urban healthcare system and did not comprehensively sample community reservoirs. Because community-associated selection may contribute to *mupA* dissemination, studies spanning healthcare and community settings will be needed to define the prevalence and spread of addicted lineages.

Overall, our results suggest that reducing mupirocin use alone is unlikely to eliminate resistance. Moreover, costly plasmid-host dependencies may evolve into a self-sustaining component of successful MRSA lineages; as with penicillin resistance [45], mupirocin resistance could become endemic. This possibility underscores the need for new decolonization strategies, such as bacteriophage-derived lysins [46], which may be less susceptible to resistance, as well as exploitation of collateral susceptibilities (e.g., isoleucine) and antibody-based approaches targeting *S. aureus* colonization. However, new interventions alone will be insufficient. Preserving their effectiveness will require more judicious use by targeting patients most likely to transmit or acquire MRSA rather than broad empiric treatment. Such risk stratification depends on genomic surveillance to identify transmission events during asymptomatic colonization, thereby identifying high-risk spreaders and enabling development of personalized models that predict transmission [12]. Because resistance may nevertheless emerge, surveillance will also be essential to detect and contain resistant clones before they adapt, spread, and become entrenched [8]. Real-time implementation of genomic surveillance could enable such early intervention [12, 47–49].

## Author Contributions

M.P., Conceptualization, Data curation, Formal analysis, Investigation, Visualization, Methodology, Writing - original draft and review and editing

A.R.W., Investigation, Writing - review and editing

S.D., Investigation, Writing - review and editing

A.T., Data curation, Writing - review and editing

G.P., Data curation, Formal analysis, Visualization, Methodology, Writing - review and editing

C.T., Data curation, Formal analysis, Visualization, Methodology, Writing - review and editing

J.M., Data curation, Writing - review and editing

S.B., Methodology, Investigation, Writing - review and editing

N.S., Data curation, Writing - review and editing

R.J.U., Conceptualization, Writing - review and editing

K.R., Data curation, Writing - review and editing

O.O., Conceptualization, Data curation, Writing - review and editing

C.O., Resources, Writing - review and editing

K.D., Conceptualization, Investigation, Writing - review and editing

M.B.O., Investigation, Resources, Writing - review and editing

A.R., Formal analysis, Conceptualization, Investigation, Writing - review and editing

A.P., Data curation, Formal analysis, Methodology, Writing - review and editing

S.H., Resources, Supervision, Funding acquisition, Writing – review and editing

B.S., Conceptualization, Resources, Supervision, Funding acquisition, Project Administration, Writing – original draft and review and editing

## Disclaimer

The funders had no role in the study design, data collection, data analysis, data interpretation, or writing of this report. The opinions, findings and conclusions expressed in this manuscript reflect those of the authors alone.

## Data availability

Sequencing data are available through the NCBI repository using the accession number PRJNA1442056.

## Acknowledgments

We thank Drs. Emily Grasso, Paul Zappile, Gael Wesby and Peter Meyn for assistance with library construction, and sequencing; Marc Lipsitch, Lorna Thorpe, and Jeffrey Weiser for critical comments on the manuscript; and Daria Frolova and John A. Lees for helpful discussions.

## Mention of any meeting(s) where the information has previously been presented

This work was presented in part at the Gordon Conference on Staphylococcal Diseases, Barcelona, Spain, August 2026.

