## Supplemental information for "Hospital transmission of methicillin-resistant *Staphylococcus aureus* driven by addictive *mupA* plasmids"

**This file includes:**

**Supplementary Information text (Methods)**

**Supplementary Figures (S1 to S7)**

**Fig. S1**. **Structural comparison of major *mupA*-containing plasmid groups identified in the surveillance cohort**

**Fig. S2**. **Dated phylogeny of the E499 *ileS1* truncation lineage.**

**Fig. S3**. Effect of mupirocin on growth of mupA-containing strains with and without chromosomal ileS1 inactivation

**Fig. S4**. **Daily colonization and transmission kinetics in the neonatal murine model.**

**Fig. S5.** Effect of the *ileS1* inactivation (‘addiction’) on colonization and transmission in a neonatal murine model

**Fig. S6. Expression of selected stringent response–associated genes in** mupA **plasmid-containing and addicted strains**

**Fig. S7.** Schematic representation of exogenous isoleucine synergizing with mupirocin to inhibit growth of *mupA*-containing strains under branched-chain amino acid (BCAA)-limited conditions.

**Supplementary Tables (S1 to S6)**

**Table S1**: Characteristics of patients harboring *mupA*-positive and *mupA*-negative *S. aureus* isolates

**Table S2**: Crude and adjusted transmission rate differences associated with *mupA* carriage

**Table S3**: Association between *mupA* carriage and transmission across MRSA clonal complexes and MSSA

**Table S4**. Bacterial strains used in the study

**Table S5**. Oligonucleotides used in the study

**Table S6**. *S. aureus* strains subjected to long-read sequencing and hybrid genome assembly for plasmid characterization.

**References for SI citations**

**Supplementary Information Text (Methods)**

**Bacterial strains, plasmids, primers and growth conditions**

*S. aureus* strains, plasmids, and primers used in the study are described in Supplementary Data (Tables 4 and 5). Bacterial cells were grown in tryptic soy broth (TSB), Luria-Bertani broth (LB; for *E.coli*) or RPMI at 37ºC or at 28 ºC with orbital shaking at 180 rpm. Colony formation was on tryptic soy agar (TSA) with or without defibrinated sheep blood, or Luria-Bertani Agar (LB) incubated at 37ºC, 30ºC or 28ºC. Where indicated, media were supplemented with chloramphenicol (15 μg/mL), mupirocin (30–250 μg/mL), cadmium chloride (0.3 mM), or isoleucine (15 μM to 10 mM).

For growth curve analysis, overnight cultures grown in TSB were diluted 1:1,000 in TSB, Roswell Park Memorial Institute medium (RPMI; Gibco, Thermo Fisher, Waltham, MA, # 11879020) or branched-chain-amino acid-deficient RPMI (US Biological, Salem, MA, #R8999). Growth was monitored at 37°C in 96-well plates (100 μL/well) with orbital shaking at 500 rpm, using BioTek LogPhase 600 microbiology reader (Agilent, Santa Clara, CA), measuring OD_600_ at 40-minute intervals. Data represent the mean of three biological replicates; each performed with at least 3 three technical replicates.

**Relacin growth assays**

Relacin (1PlusChem LLC, San Diego, CA) was dissolved in DMSO to prepare a 500 mM stock solution. Overnight cultures were diluted 1:1000 into TSB with or without relacin (8 mM). Growth was monitored at 37°C in 96-well plates (100 μL per well) using a BioTek LogPhase 600 microbiology reader (Agilent, Santa Clara, CA, USA). Optical density (OD_600_) was measured at 20-min intervals. Growth curves were performed using wild-type, plasmid-carrying (pMP1), and addicted (ileS1 Q299*) strains under identical conditions.

**Isoleucine-mediated sensitization assays**

To assess whether exogenous isoleucine altered susceptibility of *mupA*-containing strains to mupirocin, overnight cultures grown in TSB were diluted 1:1000 into branched-chain amino acid–deficient RPMI medium (US Biological, #R8999) supplemented with mupirocin (100 μg/mL) and/or isoleucine (15 or 30 μM). Branched-chain amino acid–deficient RPMI without supplementation was used for growth-control wells. Growth was monitored at 37°C in 96-well plates (100 μL per well) with orbital shaking (500 rpm) using a BioTek LogPhase 600 microbiology reader (Agilent). Optical density at 600 nm (OD_600_) was measured at 40-min intervals.

**Antimicrobials and chemicals**

Antimicrobials, chemicals, and reagents were obtained from MilliporeSigma (Burlington, MA), Thermo Fisher Scientific (Waltham, MA), US Biological (Salem, MA), New England Biolabs (NEB; Ipswich, MA), 1PlusChem LLC (San Diego, CA) and Biomerieux (Durham, NC).

**Bacterial disinfectant killing assays**

To measure lethal action, overnight cultures were diluted (OD_600_ ∼ 0.05) in fresh TSB medium and grown with shaking to early exponential (OD_600_ ∼ 0.15) phase, and treated with 20% ethyl alcohol (EtOH), 13% 2-propanol, or 20 mM H_2_O_2_ for the times indicated. At the end of treatment, aliquots were removed, concentrated by centrifugation, serially diluted in phosphate-buffered saline, and plated for determination of viable counts at 24 h. The percent survival was calculated relative to a sample taken at the time of addition of the killing agent.

**Construction of mutants**

*mupA* plasmid transconjugants

*mupA* plasmid pMP1 was transferred from clinical isolate 1394 into laboratory strains LAC Cd^R^ (BS1158), RN4220 + SaPIbov1 (BS1142), and JH1 + pOS1-GFP (BS1951) by conjugation as described.^1^ Recipient strains carried antibiotic resistance markers for counter-selection. Following conjugation, donor–recipient mixtures were plated on TSA containing mupirocin and tetracycline (30 μg/mL and 5 μg/mL, respectively) to select RN4220 transconjugants, or mupirocin and chloramphenicol (30 μg/mL and 15 μg/mL, respectively) to select JH1 transconjugants. Direct transfer from the donor strain 1394 to LAC Cd^R^ (BS1158) did not yield detectable transconjugants; therefore, RN4220 + pMP1 was used as an intermediate donor for LAC Cd^R^, with selection on mupirocin (30 μg/mL) and CdCl₂ (0·3 mM).

The resulting strains, LAC CdR + pMP1 (BS1963) and JH1 + pMP1 (BS2045), were verified by antimicrobial susceptibility testing using Etest gradient strips (BioMérieux) as described.^2^ Strain identities and plasmid content were subsequently confirmed by Oxford Nanopore and Illumina hybrid genome assembly (Plasmidsaurus, Eugene, OR). In BS2045, the multicopy GFP plasmid pOS1 was cured by a single passage on non-selective TSA, followed by screening for loss of GFP fluorescence and chloramphenicol resistance. The resulting cured derivative was re-sequenced by hybrid genome assembly to confirm plasmid loss and the absence of adventitious mutations.

ileS1 Q299 addiction mutant

The *ileS1* Q299* mutant (BS2039; pMP1 ileS1 Q299*) was constructed in the LAC derivative BS1963 carrying the native *mupA* plasmid (pMP1) using allelic replacement with pIMAY-Z, as described by Monk and Stinear.^3^ pIMAY-Z contains a thermosensitive replicon, chloramphenicol resistance marker, and lacZ reporter for blue-white screening. A 1,103-bp *ileS1* fragment carrying the Q299* (STOP) mutation was amplified from clinical isolate 52_054 and cloned into pIMAY-Z using BamHI and EcoRI restriction sites. The resulting construct was introduced into BS1963, and allelic exchange was performed according to the published protocol without modification.^3^

**Measurement of *mupA* plasmid transfer frequency**

Conjugation assays were performed as described.^4^ LAC + pMP1 and LAC + pMP1 *ileS1* Q299* served as donors, and a chloramphenicol-resistant LAC derivative served as the recipient. Transconjugants were selected on TSA containing chloramphenicol and mupirocin. Conjugation frequency was calculated as the ratio of transconjugant to donor CFU. Each donor strain was tested in five biological replicates.

**Measurement of biofilm formation**

Biofilm formation was quantified using a crystal violet assay as previously described.^5^ Twelve-well tissue culture-treated polystyrene plates (Corning, Corning, NY, #CLS3513) were coated with fibrinogen (one mL per well, 100 µg/mL) and incubated overnight. Overnight broth cultures were diluted 1:100 into fresh TSB supplemented with 1% (w/v) glucose and 3% (w/v) NaCl (TSBG), added to 12-well plates (2 mL/well), and incubated statically at 37°C for 24 h. Following incubation, supernatants were discarded and adherent biofilms were washed three times with sterile phosphate-buffered saline (PBS), heat-fixed at 60°C for 1-h and stained with 1% (w/v) crystal violet (200 µL per well) for 10 min at room temperature. Excess stain was removed, and the biofilms were washed three times with PBS. Bound crystal violet was eluted with 2 mL of 33% (v/v) acetic acid incubated for ten min, diluted 1:2 in PBS, transferred to a 96-well flat-bottom, non–tissue culture–treated polystyrene plate (Corning, #CLS3370), and absorbance was measured at 595 nm using

a Synergy Neo2 plate reader (Agilent).

**Measurement of gene expression by qRT-PCR**

Overnight broth cultures of each bacterial strain were diluted (1:100) into fresh TSB medium supplemented with 1% w/v glucose (TSBG) 3% w/v NaCl, aliquoted into 12-well (2 mL per well) tissue-culture treated polystyrene plates (Corning, #CLS3799), and incubated statically at 37°C for 6 h or 12 h. Supernatants were discarded and biofilms were resuspended in one volume (333 μL) of sterile PBS and added to a 1·5 mL tube containing two volumes (667 μL) of RNAprotect Bacteria Reagent (Qiagen, Hilden, Germany, #76506). Samples were vortexed for 5 seconds, incubated for 5 min at room temperature, then centrifuged at 5,000 x g at 4^o^C for 10 minutes.

Gene expression analysis was performed by qRT-PCR as described.^6^ Briefly, RNA was purified from biofilm-grown cells, and cDNA was synthesized using the Maxima First Strand cDNA Synthesis Kit (Thermo Fisher Scientific). Quantitative reverse-transcription PCR was performed using the QuantiNova™ SYBR Green PCR Kit (Qiagen). Primers were synthesized by IDT (Coralville, IA). Three independent biological replicates were analyzed in technical triplicate. Expression levels were normalized to rpoB, and relative gene expression was calculated using the 2^–∆∆Ct^ method.^7^

**Measurement of *S. aureus* colonization and transmission in mice**

Neonatal mouse colonization and transmission experiments were performed using a previously described model.^8^ Briefly, pups (four to seven days of age) were intranasally inoculated with 10⁶ CFU of S. aureus (LAC Cd^R^, LAC Cd^R^ + pMP1, LAC Cd^R^ + pMP1 *ileS1* Q299*, JH1, JH1 + pMP1; for strain numbers see Table S4) in three μL sterile phosphate-buffered saline (PBS) without anesthesia to restrict inoculation to the upper respiratory tract (URT). Inoculated (index) pups were immediately returned to their litter and remained cohoused with dam and littermates for the duration of the experiment. Bacterial shedding was assessed daily by gently tapping the nares of each pup 20 times onto CHROMagar plates, which were incubated at 37°C for 24–48 h to detect colonies. Transmission was assessed in contact littermates on days one to seven post-inoculation. At the experimental endpoint, pups and dam were euthanized and URT colonization was quantified by retrograde lavage. Briefly, 300 μL sterile PBS was instilled into the trachea and recovered through the nares. Lavage fluid was serially diluted in sterile PBS and plated on CHROMagar for colony-forming unit (CFU) enumeration. Index-to-contact pup ratios ranged from 1:3 to 1:4. All animal procedures were approved by the NYU Langone Health Institutional Animal Care and Use Committee (IACUC protocol # IA16-01941).

**Whole-genome sequencing and analysis**

Whole-genome sequencing, assembly, quality control, genotyping, phylogenetic analyses, and transmission inference were performed using methods previously described for the institutional genomic surveillance program.^9,10^ Briefly, genomic DNA was extracted using MagMAX DNA Multi-Sample Ultra 2.0 (Thermo Fisher Scientific), sequenced on an Illumina platform, and assembled following established quality-control procedures. Genomic analyses included assignment of sequence type and clonal complex, detection of *mecA* and SCC*mec* type, identification of Panton-Valentine leukocidin (PVL) and arginine catabolic mobile element (ACME) loci, phylogenetic reconstruction, and genomic surveillance-based transmission analyses. Additional details regarding read processing, assembly, contamination screening, genotyping, variant calling, phylogenetic reconstruction, and transmission analyses are provided in a prior publication.^9,10^

For the present study, all clinical isolates except PSCP1394 and its derivative constructs underwent Illumina sequencing. To characterize *mupA* plasmids, a subset of 27 isolates (Supplementary Table 3) underwent additional Oxford Nanopore sequencing (Plasmidsaurus, Eugene, OR). Hybrid assemblies were generated by polishing Nanopore assemblies with Illumina reads using pypolca v0.3.1 (“--careful” mode). Strain PSCP1394, a clinical isolate carrying the *mupA* plasmid used in plasmid transfer and isogenic experiments, and its derivative constructs (LAC + pMP1 [BS1963] and LAC + pMP1 *ileS1* Q299* [BS2039]) also underwent Oxford Nanopore and Illumina sequencing with hybrid assembly. Hybrid assemblies were used for plasmid characterization, assignment of plasmid structural groups, and comparative analyses of *mupA*-containing plasmids.

Assignment of *mupA* plasmid groups and distribution among S. aureus isolates

Complete *mupA* plasmid sequences were obtained from 27 isolates (Supplementary Table 6) subjected to Oxford Nanopore and Illumina hybrid assembly. Hybrid assemblies from these representative isolates, together with all short-read assemblies, were processed using MOB-recon v. 3.1.9^11^ with default parameters to assign plasmid groups and annotate characteristic plasmid genes. Plasmid contigs from hybrid assemblies were compared using minimap v. 2.28^12^ with parameters ‘-X -N 50 -p 0.1 -c’ and plotted with gggenomes (<https://thackl.github.io/gggenomes/>; Supplementary Fig. 1).

Because plasmid sequences from short-read assemblies are frequently fragmented across multiple contigs, *mupA*-encoding sequences were often located on small contigs and plasmids reconstructed by MOB-suite were not always complete. Consequently, *mupA* could not always be assigned to a reconstructed plasmid in the surveillance cohort. To maximize specificity, plasmid group assignments were restricted to assemblies in which *mupA* and adjacent plasmid group-defining sequences were present on the same contig and supported by MOB-suite predictions. This approach enabled inference of plasmid group membership for isolates lacking complete plasmid assemblies while minimizing misclassification arising from fragmented assemblies. Assigned plasmid groups were used to estimate the prevalence and distribution of *mupA* plasmids across the surveillance cohort and to annotate chromosomal phylogenies (Figs. 1A and 3A). For isolates in which *mupA* carriage could not be directly assigned to a plasmid or the chromosome, its location was inferred by assigning it to the reconstructed plasmid whose plasmid group matched that of the *mupA*-carrying plasmid in the genetically closest isolate, as determined by core-genome SNP distance.

Chromosomal integration of *mupA*

We analyzed 31 complete S. aureus genome assemblies from our historical collection (see Methods), which is enriched for mupA-positive CC8 isolates. The presence of mupA (NCBI accession NG_048008.1) was assessed using nucleotide BLAST^13^ with minimum thresholds of 95% nucleotide identity and 95% query coverage. Detection of mupA within a contig ≥2 Mb in length was interpreted as chromosomal integration. To identify mupA-addicted isolates, the 31 complete genomes were aligned to the CC8 reference genome of S. aureus strain FPR3757 (NCBI accession GCF_000013465.1) using Snippy,^14^ and variants were called in the chromosomal ileS1 (locus tag SAUSA300_RS05890). Isolates harboring stop-gain or frameshift mutations in ileS1 were classified as mupA-addicted.

**Detection of patient-to-patient S. aureus hospital transmission**

Hospital transmission events were identified using the genomic surveillance framework previously described,^9,10^ which was adapted from published genomic epidemiology approaches integrating genomic and epidemiologic data to infer transmission.^15,16^ Briefly, isolate pairs differing by ≤20 single-nucleotide variants (SNVs) were evaluated for epidemiologic linkage using electronic health record data. The 20-SNV threshold was selected on the basis of prior analyses demonstrating that within-host diversity and epidemiologically linked transmission pairs were almost uniformly separated by ≤20 SNVs. Transmission events were defined as genetically related isolates (≤20 SNVs) with epidemiologic linkage, including either direct linkage (shared unit, room, provider, or procedure during overlapping admissions) or indirect linkage (shared ward within six months without overlapping admissions). Genomically linked patients were grouped into genomic clusters defined as connected components of ≥2 patients linked by at least one ≤20-SNV comparison. Transmission clusters were defined as genomic clusters involving ≥3 patients that contained at least one epidemiologically supported transmission event. F**or all analyses, the outcome was patient-level involvement in at least one transmission event. Accordingly, transmission rates represent the proportion of patients involved in transmission rather than the number of pairwise transmission events. Because both members of a linked pair are counted as having participated in transmission, patient-level transmission rates generally exceed the number of unique pairwise transmissions.** Additional details regarding genomic thresholds, linkage criteria, and cluster construction are described in the prior publications.^9,10^

**Statistical analysis**

Prism software (GraphPad, Boston, MA) was used to perform statistical analyses. Statistical significance was determined using the Student’s *t*-test, Mann–Whitney *U* test, and ordinary one-way ANOVA, depending on the data type. Statistical significance was considered to be represented by *p* values of <0·05.

Propensity score weighting and estimation of adjusted transmission risk differences

To calculate adjusted risk ratios for transmission, we first developed a propensity score based on a logistic regression model regressing *mupA* presence/absence on factors previously identified as predictors of S. aureus transmission in our healthcare system,^10^ including lineage (MSSA vs. MRSA CC5 vs. MRSA CC8 vs. MRSA other), age, sex, race/ethnicity, Charlson score, hospital, number of hospital admissions in previous 6 months, total hospital LOS in previous 6 months, and number of antibiotic prescriptions in the previous 6 months. To identify the optimal model, we fit several models with the same covariates but with varying flexibility (including splines, interactions) and selected the model with the lowest Akaike’s Information Criterion. The final model included 3-degree of freedom natural cubic splines for all continuous variables and interactions between sex and lineage with all other variables. We used this propensity score model to create inverse probability of treatment weights as the inverse of the predicted probability of the *mupA* status of each isolate. Adjusted risk differences for transmission were calculated using an identity-link generalized linear model with robust standard errors, weighted by the inverse probability of treatment weights. Crude risk differences were calculated using the same model but without weights.

**Supplementary Figures**

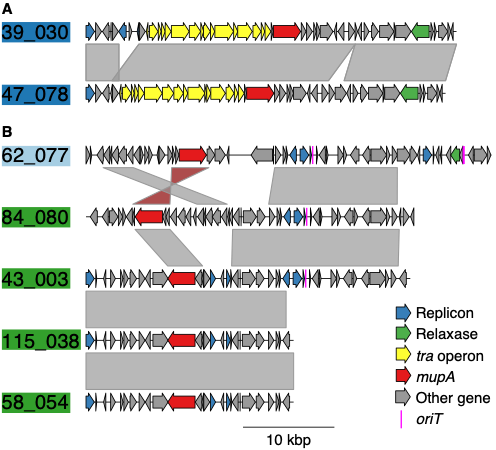

**Supplementary Figure 1. Structural comparison of representative plasmids from the three major *mupA*-associated plasmid groups.** Representative complete plasmids from the three major *mupA*-associated plasmid groups identified in the surveillance cohort were selected from 27 isolates subjected to Oxford Nanopore long-read sequencing and hybrid genome assembly for plasmid characterization (Supplementary Table 6). **The distribution of all plasmid groups across the surveillance phylogeny is shown in Fig. 1A and, among** mupA**-positive isolates, in Fig. 3B.** Plasmids are grouped according to structural similarity. Mobility predictions corresponded closely with the major structural groups, consistent with a recent regional survey of complete *mupA*-containing plasmids.^17^ **(A)** Representative Group 1 plasmids (39_030, 47_078; dark blue), predicted to be conjugative. **(B)** Representative Group 2 plasmids (green), including both mobilizable (84_080, 43_00) and non-mobilizable (115_038, 58_054) examples, and a representative Group 3 plasmid (67_077; light blue), predicted to be mobilizable. Plasmids were compared using minimap2.^12^ Coding sequences (CDSs) are shown as arrows and colored according to predicted function. ***mupA* (red)**, replicon (rep) genes (blue), conjugation-associated genes (yellow), and origin(s) of transfer (oriT; pink) are highlighted; other genes are shown in gray. Plasmids are ordered to emphasize structural homology within and between groups. Shaded regions between plasmids indicate homologous nucleotide sequences identified by pairwise alignment, with red shading indicating an inversion.

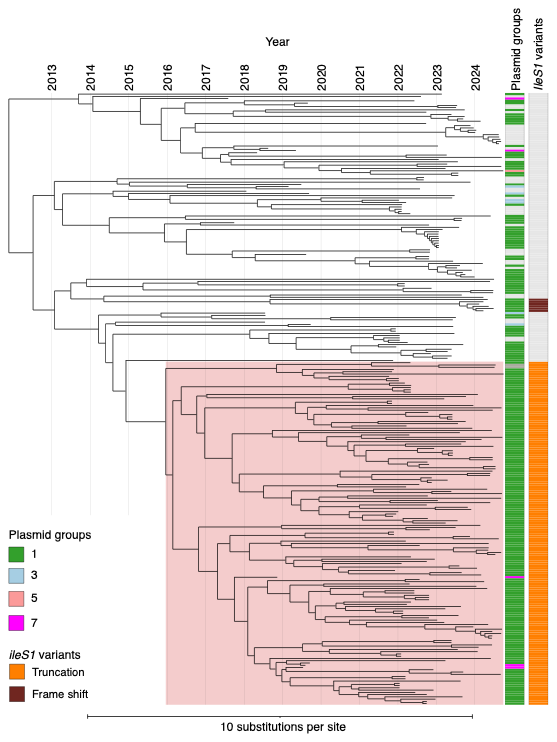

**Supplementary Figure 2. Dated phylogeny of the E499 *ileS1* truncation lineage.** Time-scaled phylogeny of isolates belonging to the E499 ileS1 truncation lineage identified among mupA-positive MRSA isolates. The E499 lineage identified in the contemporary surveillance cohort (n = 271 isolates) is highlighted. To reconstruct the evolutionary history of the lineage, phylogenetically related isolates from historical collections (2017–2020) and the contemporary surveillance cohort (2022–2025) were included in the analysis. Annotation tracks indicate year of isolation, Mob-suite-imputed plasmid group, and ileS1 inactivation status. The dated phylogeny supports emergence of the E499 lineage before the start of genomic surveillance, with an inferred common ancestor dating to approximately 2015–2016. The lineage includes both mupA-positive and mupA-negative descendants derived from a common ancestor.

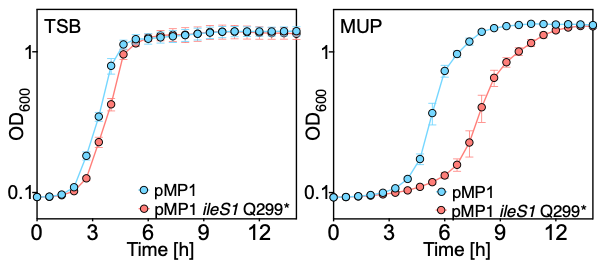

**Supplemental Fig. 3.** **Effect of mupirocin on growth of mupA-containing strains with and without chromosomal ileS1 inactivation. (A)** Overnight cultures of S. aureus LAC (BS1158) carrying the mupA-containing plasmid pMP1 (BS1963) or an isogenic ileS1 truncation mutant (ileS1 Q299* [BS2039]) were diluted 1000 x in fresh tryptic soy broth (TSB) without antibiotic. Growth was monitored by measuring the optical density at 600 nm (OD_600_). **(B)** Growth of the same strains in TSB as in A supplemented with mupirocin (250 μg/mL). Mupirocin MICs were unchanged in the ileS1 Q299* mutant relative to the parental strain. Data represent the means ± SD from biological replicates (*n* = 3). Thus, addiction does not enhance resistance or fitness in the presence of mupirocin.

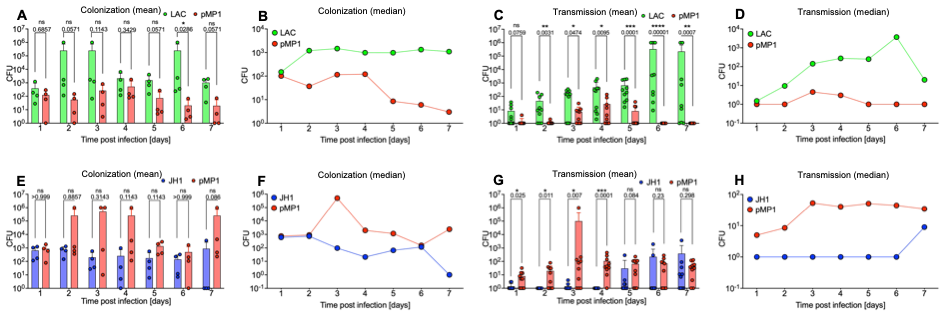

**Supplementary Fig. 4. Daily colonization and transmission kinetics in the neonatal murine model.** Neonatal mice were inoculated and monitored as described in Fig. 5. **(A–D)** Colonization and transmission kinetics of the CC8 strain LAC and its pMP1-containing derivative (BS1158 and BS1963, respectively). **(E–H)** Colonization and transmission kinetics of the CC5 strain JH1 and its pMP1-containing derivative (BS1231 and BS2045, respectively.) **(A, B, E, F)** Daily nasal colonization burden among inoculated index pups. **(C, D, G, H)** Daily nasal colonization burden among uninoculated contact pups following transmission. **(A, C, E, G)** Mean CFU at each time point. **(B, D, F, H)** Median CFU at each time point. All y-axes are shown as a logarithmic scale. To enable plotting on the log scale, a value of 1 was assigned to samples with no detectable S. aureus growth. Multiple Mann-Whitney tests were used to determine statistical differences between samples (**p*<0.05, ***p*<0.01; ****p*<0.001; *****p*<0.0001) in panel A, C, E and G. Consistent with the cumulative analyses shown in Fig. 5, pMP1 reduced colonization and transmission in CC8 but increased both measures in CC5, indicating that the fitness consequences of plasmid carriage are strain dependent.

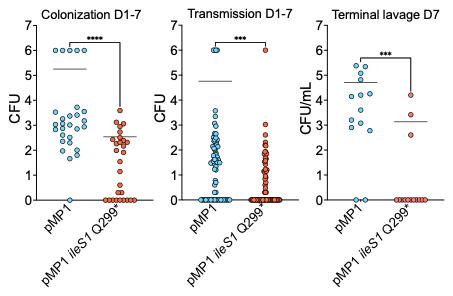

**Supplementary Fig. 5. Effect of the *ileS1* inactivation (‘addiction’) on colonization and transmission in a neonatal murine model.** Colonization and transmission of the CC8 strain LAC + pMP1 (BS1963, labeled here as pMP1 for simplicity) and LAC + pMP1 *ileS1* Q299* (BS2039, labeled as pMP1 *ileS1* Q299*); Left, cumulative nasal colonization burden among inoculated index pups measured over days 1–7 post-inoculation. Middle, cumulative transmission burden among uninoculated contact pups measured over the same interval. Right, terminal upper respiratory tract colonization quantified by retrograde nasal lavage at the experimental endpoint. In each panel, blue indicates the *mupA*-plasmid–containing strain with chromosomal *ileS1* inactivation. Each symbol represents an individual animal. All y-axes are shown on a logarithmic scale. A value of 1 was assigned to samples with no detectable S. aureus growth. To enable plotting on the log scale, a value of 1 was assigned to animals from which no S. aureus colony was recovered.

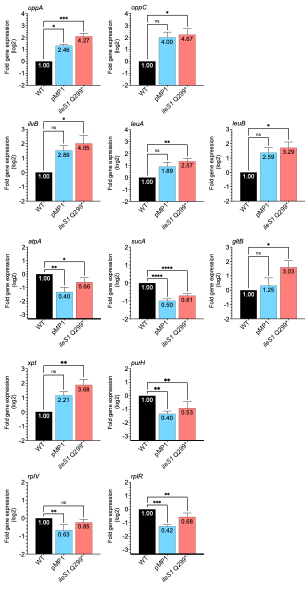

**Supplementary Fig. 6. Expression of selected stringent response-associated genes in mupA plasmid-containing and addicted strains.** Total cellular RNA was extracted from 12-h biofilm growth of S. aureus LAC wild-type (WT; BS1158), the isogenic mupA plasmid-containing derivative (pMP1; BS1963), and the addicted pMP1 ileS1 Q299* mutant (BS2039). Transcript abundance was quantified by reverse transcription quantitative PCR using rpoB as an internal control. Genes were selected to represent functional pathways associated with the stringent response, including oligopeptide transport, amino acid biosynthesis, central metabolism, and purine biosynthesis. Expression levels were normalized to those of the wild-type, which was assigned a value of 1. Bars indicate fold-change relative to wild type. Data represent the mean ± SEM of three independent experiments. Ordinary one-way Anova was used to determine statistical differences between samples (**p*<0.05, ***p*<0.01; ****p*<0.001; *****p*<0.0001). The addicted strain generally showed larger effect sizes across several markers, with the clearest differences involving branched-chain amino acid biosynthesis genes (ilvB, leuA, and leuB), consistent with stronger stringent response-mediated relief of CodY repression.^11,12^ These transcriptional changes parallel the enhanced biofilm phenotype observed in addicted strains, a phenotype previously associated with CodY regulation.^18^

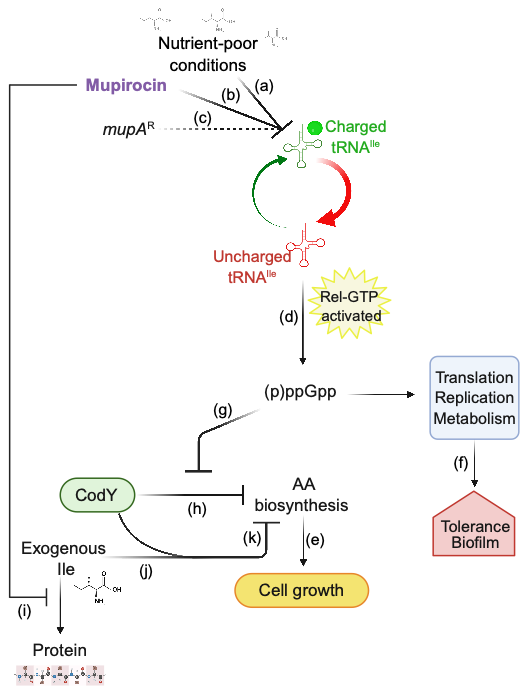

**Supplementary Fig. 7. Schematic description of exogenous isoleucine synergizing with mupirocin to inhibit growth of *mupA*-containing strains under branched-chain amino acid (BCAA)-limited conditions.** Under nutrient-poor conditions with limited BCAA availability (*a*), mupirocin partially inhibits isoleucyl-tRNA charging (*b*) despite the presence of the plasmid-encoded *mupA* resistance determinant (*c*). This shifts the balance from charged to uncharged tRNA-Ile, resulting in accumulation of uncharged tRNA-Ile at the ribosomal A site and low-level activation of the Rel-dependent stringent response. Activation of the stringent response increases production of the alarmone ppGpp (*d*), which suppresses translation, replication, and growth-associated metabolism (*e*) while promoting stress tolerance and biofilm formation (*f*). In parallel, ppGpp antagonizes CodY (*g*), relieving repression of amino acid (AA) biosynthetic pathways (*h*) and promoting adaptation to nutrient limitation and continued protein synthesis (*i*). Thus, cells can continue to grow despite ongoing mupirocin exposure. In this scenario, addition of exogenous isoleucine disrupts adaptation by maintaining CodY activity (*j*), thereby preventing induction of biosynthetic pathways required to replenish limiting amino acids (*k*). Furthermore, we hypothesize that the growth-inhibitory effect of exogenous isoleucine is amplified by mupirocin. By impairing incorporation of isoleucine into protein synthesis, mupirocin is expected to increase the effective intracellular availability of isoleucine or dysregulate intracellular isoleucine pools (*l*), allowing exogenous isoleucine to exert a disproportionate effect on intracellular isoleucine sensing and CodY activation. Under these conditions, even low concentrations of exogenous isoleucine, which are minimally growth inhibitory on their own, may be sufficient to maintain CodY activity despite persistent stringent-response signaling. This regulatory mismatch between amino acid sensing and the metabolic demands imposed by BCAA limitation prevents appropriate induction of amino acid biosynthetic pathways, creating a conditional vulnerability in *mupA*-containing strains whereby exogenous isoleucine and mupirocin synergize (*h* and *k*), culminating in growth collapse under BCAA-limited conditions.

**Supplementary Tables**

**Supplementary Table 1. Characteristics of patients harboring *mupA*-positive and *mupA*-negative *S. aureus* isolates.**

|  | *mupA* (-)  (*n* = 9,108) | *mupA* (+)  (*n* = 1,213) |
| --- | --- | --- |
| Age in years - mean (SD) | 62 (19) | 65 (18) |
| Age group - n (%) |  |  |
| 18-24 | 200 (2.2%) | 19 (1.6%) |
| 25-34 | 648 (7.1%) | 75 (6.2%) |
| 35-44 | 1,012 (11%) | 91 (7.5%) |
| 45-54 | 1,061 (12%) | 137 (11%) |
| 55-64 | 1,554 (17%) | 192 (16%) |
| 65-74 | 1,794 (20%) | 261 (22%) |
| 75+ | 2,839 (31%) | 438 (36%) |
| Gender |  |  |
| Male | 5,369 (59%) | 771 (64%) |
| Female | 3,732 (41%) | 442 (36%) |
| Missing | 7 | 0 |
| Race/ethnicity |  |  |
| Asian | 787 (9.2%) | 39 (3.4%) |
| Black | 1,309 (15%) | 124 (11%) |
| Hispanic | 1,996 (23%) | 186 (16%) |
| Other | 76 (0.9%) | 6 (0.5%) |
| White | 4,424 (51%) | 791 (69%) |
| Missing | 516 | 67 |
| Charlson index score |  |  |
| 0 | 4,585 (50%) | 603 (50%) |
| 1-2 | 2,162 (24%) | 266 (22%) |
| 3-4 | 1,556 (17%) | 206 (17%) |
| 5+ | 805 (8.8%) | 138 (11%) |
| Hospital |  |  |
| Brooklyn | 2,306 (25%) | 262 (22%) |
| Tisch | 6,802 (75%) | 951 (78%) |
| # of Hospital Admissions | 1.73 (3.19) | 2.25 (3.28) |
| Cumulative Hospital Days | 8 (19) | 15 (26) |
| Cumulative # of Antibiotics | 10 (34) | 23 (51) |
| *S. aureus* type |  |  |
| MRSA | 3,088 (34%) | 1,041 (86%) |
| MSSA | 6,020 (66%) | 172 (14%) |
| Clonal complex |  |  |
| CC5 | 2,035 (22%) | 227 (19%) |
| CC8 | 2,329 (26%) | 859 (71%) |
| Other | 4,744 (52%) | 127 (10%) |
| Involved in Transmission Event | 467 (5.1%) | 190 (16%) |

Data are presented as mean (SD) or n (%).

Abbreviations: MRSA, methicillin-resistant Staphylococcus aureus; MSSA, methicillin-susceptible S. aureus; CC, clonal complex.

**Supplementary Table 2. Crude and adjusted transmission rate differences associated with *mupA* carriage.**

|  | Rate difference (95% CI) |
| --- | --- |
| Crude | 10.5 (8.4, 12.7) |
| Adjusted^a^ | 11.3 (5.8, 16.9) |

^a^Adjusted using inverse probability of treatment weighting (IPTW) for MRSA versus MSSA status, clonal complex, hospitalization history (number of prior hospital admissions, cumulative hospital days), cumulative antibiotic exposure, age, sex, race/ethnicity, and Charlson comorbidity score.

**Supplementary Table 3. Association between *mupA* carriage and transmission across MRSA clonal complexes and MSSA.**

|  | **MRSA CC5** | | | | **MRSA CC8** | | | | **MRSA Other CC** | | | **MSSA** | | |
| --- | --- | --- | --- | --- | --- | --- | --- | --- | --- | --- | --- | --- | --- | --- |
|  | ***mupA* (+)** | ***mupA* (-)** | **RD** | ***mupA* (+)** | | ***mupA* (-)** | **RD** | ***mupA* (+)** | | ***mupA* (-)** | **RD** | ***mupA* (+)** | ***mupA* (-)** | **RD** |
| **Transmission** | 51 (26.3%) | 135 (12.9%) | 13.4 (6.1, 20.7) | 119 (14.6%) | | 109 (7.4%) | 7.3 (4.5, 10.1) | 8 (23.5%) | | 95 (17.0%) | 6.6 (-5.6, 18.7) | 12 (7.0%) | 128 (2.1%) | 4.9 (1.0, 8.7) |
| **No transmission** | 143 (73.7%) | 913 (87.1%) |  | 694 (85.4%) | | 1371 (92.6%) |  | 26 (76.5%) | | 465 (83.0%) |  | 160 (93.0%) | 5892 (97.9%) |  |
| **Total** | 194 | 1048 |  | 813 | | 1480 |  | 34 | | 560 |  | 172 | 6020 |  |

Data are presented as n (%) of isolates within each lineage group stratified by *mupA* carriage and transmission status. Rate differences (RDs) compare the rate of transmission among *mupA*-positive versus *mupA*-negative isolates within each lineage group; 95% confidence intervals are shown in parentheses. Because CC5 and CC8 accounted for approximately 86% of MRSA isolates in the cohort, conclusions regarding lineage-specific effects in MRSA were based primarily on these clonal complexes.

Abbreviations: MRSA, methicillin-resistant Staphylococcus aureus; MSSA, methicillin-susceptible S. aureus; CC, clonal complex.

**Supplementary Table 4. Bacterial strains^a^.**

| Strain | Background | Relevant Genotype | Reference or Source |
| --- | --- | --- | --- |
| BS1158 | LAC, USA300, CA-MRSA | *agr* group I wild-type (CC8), Erm^S^, CadA in SaPI1 *attS* | 19 |
| 1394 | *S. aureus* clinical isolate | CC5 MRSA mupR, harboring a native pMP1 plasmid | This study |
| BS1963 | BS1158 | LAC Cd^R^ + pMP1 | This study |
| BS2039 | BS1158 | LAC Cd^R^ + pMP1 *ileS1* Q299* | This study |
| BS1231 | JH1 | *agr* group II wild-type (CC5), | 20 |
| BS2045 | JH1 | JH1 WT + pMP1 | 21 |
| BS1462 | *E. coli* DH5α | DH5α with pIMAY-Z (Cm^R^ in *E. coli*; Cm^R^ in *S. aureus*) | 22 |
| BS1000 | *E. coli* IM08B | SA08BΩP_N25_-hsdS (CC8-1) (SAUSA300_0406) of NRS384 integrated between the essQ and cspB genes | 22 |
| BS1142 | RN4220 | SaPIbov1::*tetM* | 23 |
| BS1958 | BS1142 | RN4220 + pMP1 | This study |
| 52_054 | *S. aureus* clinical isolate | CC8 MRSA mup^R^ *ileS1* mutated at 895nt C>T, Q299STOP | This study |
| BS1951 | BS1231 | pOS1-GFP empty vector | This study |

^a^All bacterial strains are *S. aureus*, unless otherwise indicated

Abbreviations: CC, clonal complex; nt, nucleotide.

**Supplementary Table 5. Oligonucleotides.**

| Name | Gene/  Target | | Sequence 5’🡪 3’ | Reference  or Source |
| --- | --- | --- | --- | --- |
| ***ileS1* cloning and introduction of the inactivating mutation** | | | | |
| *Fragment.FOR ileS1 mut* | *ileS1* | | CCCCCCCTCGAGGTCGACGGTATCGATAAGCTTGATATCGGAATTTGCTTTAGAACAAATTGAATTACAGAAAAAAG | This study |
| *Fragment.REV ileS1 mut* |  |  | TTGGAGCTCCACCGCGGTGGCGGCCGCTCTAGAACTAGTGTTTGAACCGTGTTCTGCAAATAAATC | This study |
| *ileS1 ver FOR* |  |  | CTTGTCTGACGCTGTAGCAGAAGCACTG | This study |
| *ileS1 ver REV* |  |  | CTTTATCATCGATTGGACTAATTACTGGC | This study |
| **qRT-PCR** | | | | |
| *rpoB F* | *rpoB* | | GAACATGCAACGTCAAGCAG | 24 |
| *rpoB R* |  |  | AATAGCCGCACCAGAATCAC |  |
| *oppA RT1* | *oppA* | | ACGATCCACAAAGTACTATTGC | 25 |
| *oppA RT2* |  |  | TAAATGCGTCATCAATGCTGTT |  |
| *oppC RT1* | *oppC* | | ATGTCTTTTTAGGATTAGCAGC |  |
| *oppC RT2* |  |  | GTCAGTACCTAGTAGATGTTGA |  |
| *ilvB RT1* | *ilvB* | | GCGTTCAGGATCAGAAGTGC | 26 |
| *ilvB RT2* |  |  | CACCTTGTTCGTGTCTTGCT |  |
| *xpt RT1* | *xpt* | | GATTACTAAAATCTTAACCATTGAAGC | This study |
| *xpt RT2* |  |  | ATGAATAGATGTTTCATAATAACCATCC |  |
| *purH RT1* | *purH* | | ATCAAGAAGTATTGACGCGATTAAG | 27 |
| *purH RT2* |  |  | GATTGTTGTGGATTTTCTCCATATC |  |
| *atpA RT1* | *atpA* | TAGAAAGAGCAGCAAAATTAAA | | 28 |
| *atpA RT2* |  | TTGTTGGTACATAAGCTGAAAT | |  |
| *sucA RT1* | *sucA* | GCCGTGTTACATGATGAGCA | | 29 |
| *sucA RT2* |  | CACCATATTGCGCTTCCCAA | |  |
| *rplR RT1* | *rplR* | CTTGCTGCTTCAGCTAATGC | | This study |
| *rplR RT2* |  | CTTCAAAAGACAGCGACATTGCTAC | |  |
| *rplV RT1* | *rplV* | CTTTAGCTTCTTCTTTACCGTCACTTAC | | This study |
| *rplV RT2* |  | GGACCAACATTAAAACGTTTCC | |  |
| *leuA RT1* | *leuA* | GTGAACGGTATTGGTGAAAGAG | | 30 |
| *leuA RT2* |  | GTGGTCCTTCCTTACATATAAAGC | |  |
| *leuB RT1* | *leuB* | GGTGCAATCGGTGGACCTAA | | 31 |
| *leuB RT2* |  | TAGCGCCTTTGACAACGGTA | |  |
| *gltB RT1* | *gltB* | TTGCGAAGACAAGAGGGT | | 32 |
| *gltB RT2* |  | CTAAACCAATCTCCCAAGGA | |  |

**Supplementary Table 6.** *S. aureus* strains subjected to long-read sequencing and hybrid genome assembly for plasmid characterization.

| # | Strain | Lineage |
| --- | --- | --- |
| 1 | 124_091 | CC121 |
| 2 | 124_098 | CC121 |
| 3 | 18_072 | CC121 |
| 4 | 128_053 | CC5 |
| 5 | 16_019 | CC5 |
| 6 | 39_030 | CC5 |
| 7 | 39_099 | CC5 |
| 8 | 42_100 | CC5 |
| 9 | 43_003 | CC5 |
| 10 | 46_001 | CC5 |
| 11 | 47_078 | CC5 |
| 12 | 58_054 | CC5 |
| 13 | 62_077 | CC5 |
| 14 | 7_079 | CC5 |
| 15 | 77_076 | CC5 |
| 16 | 1394 | CC5 |
| 17 | 101_092 | CC8 |
| 18 | 11_087 | CC8 |
| 19 | 112_052 | CC8 |
| 20 | 115_038 | CC8 |
| 21 | 25_073 | CC8 |
| 22 | 26_002 | CC8 |
| 23 | 28_003 | CC8 |
| 24 | 3_092 | CC8 |
| 25 | 49_077 | CC8 |
| 26 | 53_075 | CC8 |
| 27 | 84_080 | CC8 |

**Note**. Strains were selected for long-read sequencing to characterize the diversity of *mupA* plasmids identified within the surveillance cohort. Complete plasmid sequences were generated using Oxford Nanopore long-read sequencing and hybrid assembly.
